# Perioperative non-invasive brain stimulation for the prevention of postoperative delirium: a systematic review and meta-analysis of randomised controlled trials

**DOI:** 10.64898/2026.11.02.26367005

**Authors:** Huaiming Wang, Hui Pan, Deshan Li, Aimin Zhang, Yanruo Huang, Xing Liu, Jinjun Shu, Hongwei Zhang, Dan Zhang, Jiang Zou, Li Du

## Abstract

**Background:** Postoperative delirium is a common perioperative complication. Early randomised trials suggested that non-invasive brain stimulation may reduce postoperative delirium, whereas the recent MODEST trial found no preventive benefit. We synthesised the randomised evidence for perioperative non-invasive brain stimulation

**Methods:** We searched eight bibliographic databases and ClinicalTrials.gov from inception to 30 September 2026 for randomised controlled trials of perioperative non-invasive brain stimulation in adults undergoing surgery. The primary outcome was postoperative delirium. Random-effects meta-analyses estimated risk ratios (RRs), mean differences, or standardised mean differences. Risk of bias was assessed using RoB 2 and certainty of evidence using GRADE.

**Results:** Nine trials contributed data from 1451 participants. Postoperative delirium occurred in 72 of 750 participants receiving active stimulation and 166 of 701 controls (RR 0.41, 95% confidence interval [CI] 0.31–0.56; *I*^2^=24.7%; moderate-certainty evidence). Estimates also favoured stimulation for hypoactive and hyperactive delirium, but not clearly for mixed delirium. Pain, anxiety, and sleep outcomes favoured stimulation but were highly heterogeneous and supported by very-low-certainty evidence. Length of hospital stay was not reduced. Adverse events could not be pooled because reporting was incomplete and inconsistent.

**Conclusions:** Perioperative non-invasive brain stimulation may reduce postoperative delirium, but the evidence does not establish a class effect or support routine clinical use. Geographical concentration, methodological concerns, heterogeneous stimulation protocols, and discordance with the MODEST trial warrant independent multicentre sham-controlled trials.

**PROSPERO registration:** CRD420261522168

## Introduction

Postoperative delirium (POD) is an acute, fluctuating disturbance of attention and cognition that occurs after surgery and anaesthesia and is associated with prolonged hospitalisation, functional decline, institutionalisation, persistent cognitive impairment, and mortality.^1, 2^ Contemporary perioperative guidelines emphasise risk assessment, validated postoperative screening, and multicomponent non-pharmacological prevention.^3-5^ Nevertheless, POD remains common, and no preventive strategy is universally effective or suitable for all surgical patients.

A recent network meta-analysis suggested that dexmedetomidine and several other pharmacological interventions may reduce POD in selected settings.^6^ However, treatment-specific adverse effects and variable certainty of evidence limit the applicability of these findings, supporting continued evaluation of mechanistically distinct non-pharmacological interventions.

Non-invasive brain stimulation (NIBS) techniques, including transcranial direct current stimulation (tDCS), transcranial alternating current stimulation (tACS), repetitive transcranial magnetic stimulation (rTMS), and continuous theta-burst stimulation, modulate cortical excitability or oscillatory activity through electrical or magnetic stimulation.^7^ These interventions may influence neural networks involved in attention, arousal, and executive function, but their effects depend on the stimulation modality, target, intensity, frequency, timing, and underlying brain state. Different NIBS modalities should therefore not be assumed to have interchangeable clinical effects.

Evidence from other POD prevention strategies also illustrates the limitations of relying on a pooled estimate alone: processed electroencephalography-guided anaesthesia has shown heterogeneous trial-level effects, and a Bayesian analysis of dexmedetomidine found that a large discordant trial materially altered the interpretation of earlier positive evidence.^8, 9^ This concern is directly relevant to the emerging NIBS literature. Since 2023, randomised trials have evaluated perioperative tDCS, tACS, rTMS, and continuous theta-burst stimulation across several surgical populations.^10-18^ Most early trials reported substantial reductions in POD, whereas the 2026 MODEST trial found no preventive benefit of either alpha-frequency tACS or tDCS and observed a numerically higher delirium incidence in the active groups than in sham controls.^18^This discordance raises questions about the reproducibility of the pooled effect, differences between stimulation modalities, and the influence of study quality and trial size. We therefore conducted a systematic review and meta-analysis of randomised controlled trials evaluating perioperative NIBS for the prevention of POD.

## Methods

### Review conduct and reporting

This systematic review and meta-analysis was conducted in accordance with PRISMA 2020 and methodological guidance in the Cochrane Handbook for Systematic Reviews of Interventions.^19, 20^ The completed PRISMA checklist and full electronic search strategies are provided in Supplementary Tables S1 and S2. The review was registered with PROSPERO (registration number CRD420261522168).

### Eligibility criteria

We included randomised controlled trials enrolling adults undergoing surgery and evaluating a perioperative non-invasive transcranial stimulation intervention intended to prevent POD. Eligible modalities included tDCS, tACS, rTMS, and theta-burst stimulation, irrespective of stimulation target, dose, or whether treatment was delivered before or after surgery. Comparators included sham stimulation or usual/conventional delirium-prevention care. Trials were required to report POD using a prespecified clinical assessment or delirium diagnostic instrument. We excluded non-randomised studies, studies in non-surgical populations, interventions delivered solely to treat established POD, protocols without outcome data, registry records without a published result report, duplicate reports, and studies without usable POD outcomes. Reports in any language were eligible.

### Information sources and search strategy

We searched PubMed, Embase, the Cochrane Central Register of Controlled Trials (CENTRAL), Web of Science, CINAHL, ClinicalTrials.gov, China National Knowledge Infrastructure (CNKI), Wanfang Data, and SinoMed from inception to 30 September 2026. Search concepts combined controlled vocabulary and free-text terms for transcranial magnetic or electrical stimulation, including TMS, rTMS, TBS, iTBS, cTBS, tDCS, tACS, tRNS, tPCS, NIBS, and related variants, with delirium and perioperative neurocognitive terms. Reference lists and trial records identified during screening were also examined for additional eligible reports. No language restriction was applied. Complete database-specific strategies and record counts are presented in Supplementary Table S2.

### Study selection and data extraction

Titles and abstracts were screened independently by HW and HP, who also independently assessed potentially eligible full-text reports. Disagreements were resolved through discussion and, when necessary, adjudicated by LD. Data were extracted independently in duplicate by four reviewer pairs (HW and HP; DL and AZ; YH and XL; JS and HZ) using a structured extraction form. Discrepancies were resolved by consensus; unresolved cases were reviewed by LD and, when necessary, adjudicated by DZ or JZ. Extracted variables included country, sample size, age, sex, surgical population, stimulation modality, target or montage, intensity or frequency, session duration, timing and number of sessions, comparator, delirium assessment method and observation window, outcome data, and adverse events. Where a report contained more than one eligible active arm sharing a common control, groups were handled to avoid double counting of the comparator in class-level analyses.^20^

### Outcomes

The primary outcome was incidence of POD during the postoperative assessment window defined by each trial. Validated instruments used across the included trials included the Confusion Assessment Method (CAM), CAM for the Intensive Care Unit (CAM-ICU), and 3-Minute Diagnostic Interview for CAM-defined Delirium (3D-CAM).^21-23^ Secondary outcomes were hypoactive, hyperactive, and mixed delirium; postoperative pain; anxiety; sleep disturbance; length of hospital stay; and adverse events. For pain, anxiety, and sleep outcomes measured repeatedly, postoperative day 1 was used for quantitative synthesis. Adverse events were pooled only if participant-level denominators and non-overlapping counts permitted estimation of the proportion experiencing at least one event.

### Risk of bias and certainty of evidence

Risk of bias for the POD outcome from each trial was assessed independently by HW and LD using the Cochrane Risk of Bias 2 (RoB 2) tool. Assessments covered bias arising from the randomisation process, deviations from intended interventions, missing outcome data, measurement of the outcome, and selection of the reported result.^24^ Disagreements were resolved by consensus or, when necessary, through consultation with DL; unresolved cases were adjudicated by DZ or JZ. Risk-of-bias visualisations were generated using robvis.^25^

Certainty of evidence was assessed independently by DL and AZ using the GRADE approach across the domains of risk of bias, inconsistency, indirectness, imprecision, and publication bias.^26-31^ Disagreements were resolved by consensus and, when necessary, adjudicated by JZ or HP. Randomised evidence started at high certainty and was downgraded when concerns in one or more domains were judged sufficiently serious. For syntheses including fewer than 10 studies, publication bias was assessed qualitatively because formal tests of funnel-plot asymmetry would have limited statistical power.

### Statistical analysis

For dichotomous outcomes, treatment effects were expressed as risk ratios (RRs) with 95% confidence intervals (CIs). For continuous outcomes measured on the same scale we used mean differences (MDs); when different instruments assessed the same construct, we used standardised mean differences (SMDs). When required, medians and dispersion measures were converted to approximate means and standard deviations using established methods.^32, 33^ For single-zero binary comparisons, a continuity correction of 0.5 was applied. Double-zero comparisons did not contribute an estimable RR.

We used inverse-variance random-effects meta-analysis because variation in surgical populations and stimulation protocols was expected a priori. Between-study variance (tau^2^) was estimated using restricted maximum likelihood; heterogeneity was described using Cochran Q, tau^2^, and I^2^.^34, 35^ Conventional random-effects inference was supplemented by a Modified Hartung-Knapp analysis because the number of contributing studies was small.^36^ Sensitivity analyses excluded Chen 2026, which used conventional multimodal delirium prevention without sham stimulation and paper-specific CAM thresholds; excluded trials judged at high overall risk of bias; and excluded the MODEST trial to quantify the influence of the recent discordant electrical-stimulation study. We also compared magnetic stimulation (rTMS/cTBS) with electrical stimulation (tDCS/tACS) in a subgroup analysis. In the MODEST multi-arm trial, the tACS and tDCS active arms were combined for the overall electrical-stimulation class comparison against the shared sham group to avoid double counting.

Small-study effects were explored visually using a funnel plot for the primary outcome. Formal regression-based tests for funnel-plot asymmetry, including Egger testing, were not performed because only nine studies were available, a setting in which these tests have low power and can be misleading.^37^ Analyses were performed in R version 4.6.1 (R Foundation for Statistical Computing, Vienna, Austria) using the meta package version 8.5-0.^38^ All tests were two-sided, and 95% CIs are reported.

## Results

### Study selection and characteristics

The search identified 508 records: 62 from PubMed, 28 from Embase, 97 from CENTRAL, 23 from CINAHL, 67 from Web of Science, 11 from CNKI, 177 from Wanfang Data, 24 from SinoMed, and 19 from ClinicalTrials.gov. After removal of 108 duplicates, 400 records were screened; 358 were excluded at title/abstract screening and 42 reports underwent full-text assessment. Thirty-three were excluded: 17 registry-only records, seven with insufficient outcome reporting, four with ineligible outcomes, two studies of POD treatment rather than prevention, one duplicate report, one ineligible study design, and one protocol without results. Nine trials were included (Figure 1; Supplementary Table S3).

**Fig 1.**
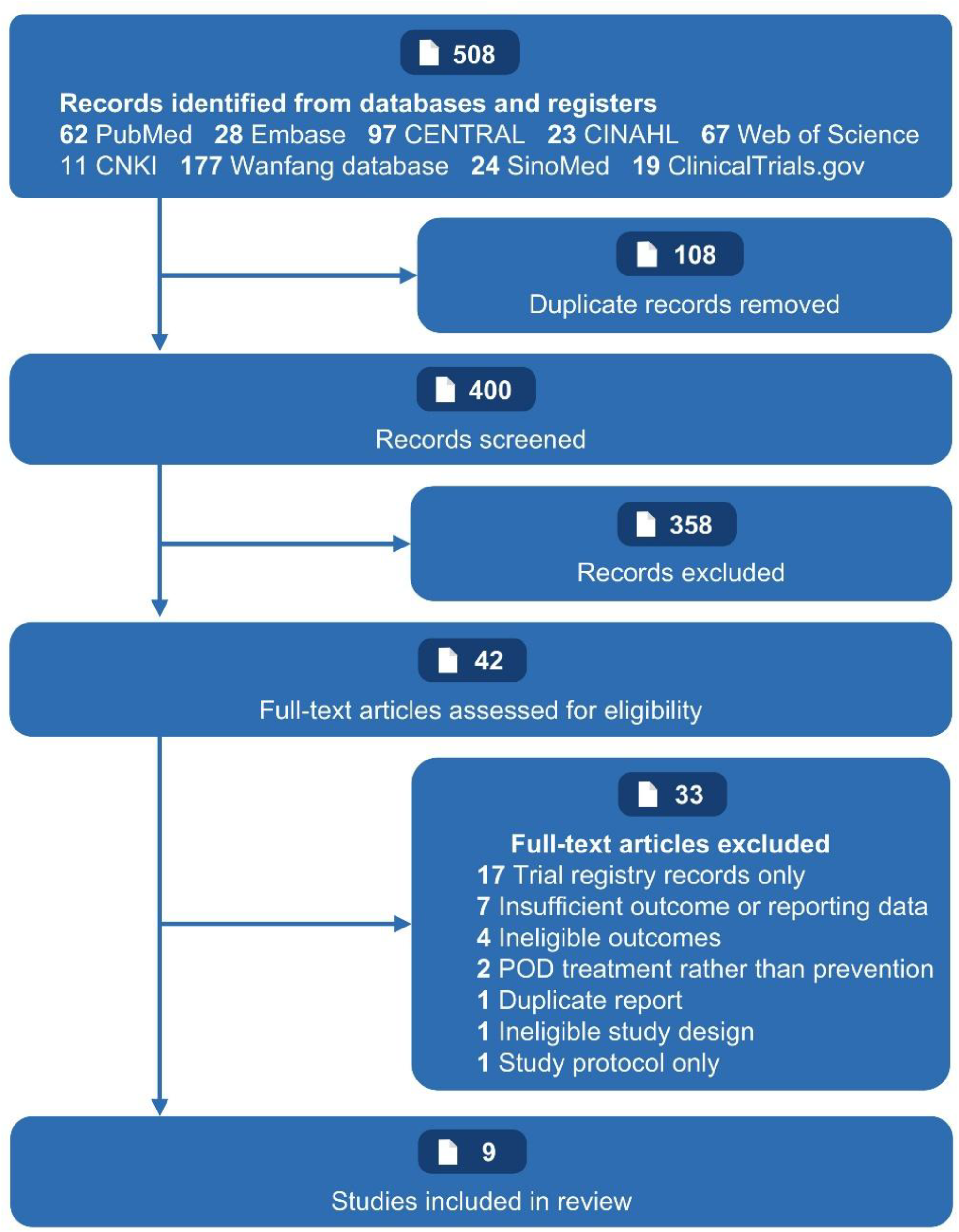
PRISMA flow diagram of study selection. The diagram shows the identification, screening, eligibility assessment, and inclusion of randomised controlled trials evaluating perioperative non-invasive brain stimulation for the prevention of postoperative delirium. PRISMA, Preferred Reporting Items for Systematic Reviews and Meta-Analyses.

The nine trials contributed 1451 participants to the principal intervention-control comparisons and were published between 2023 and 2026.^10-18^ Eight trials were conducted in China and one in Germany. Sample sizes ranged from 78 to 249. Surgical populations included lower-limb arthroplasty, colorectal cancer surgery, major abdominal surgery, prolonged laparoscopic surgery, elective non-cardiac surgery, abdominal tumour surgery, cardiac valve surgery, mixed major surgery, and hip-fracture surgery (Table 1). Four trials evaluated tDCS alone, three evaluated rTMS, one evaluated cTBS, and the MODEST trial evaluated separate alpha-tACS and tDCS active arms against a shared sham group.^10-18^ Stimulation targets, intensities, timing, and numbers of sessions varied substantially; most magnetic and tDCS protocols targeted the left dorsolateral prefrontal cortex, whereas cTBS targeted the posterior cingulate cortex and alpha-tACS targeted posterior parietal regions (Supplementary Table S4). POD was assessed using CAM, CAM-ICU, 3D-CAM, or paper-specific CAM thresholds over windows ranging from postoperative days 1-3 to 7 days or hospital discharge (Supplementary Table S5).

**Table 1.** Characteristics of the included studies.

| **Study** | **Country** | **Sample size, n** | **Female, n (%)** | **Age, mean (SD)** | **Surgical procedure** | **Stimulation modality** | **Intervention timing** | **POD assessment** |
|---|---|---|---|---|---|---|---|---|
| **Tao 2023** | China | 122 | 80(65.6) | 71.0(6.1)* | Total hip or knee arthroplasty | tDCS | Immediately after extubation in PACU | CAM/CAM-ICU; POD1–3 |
| **Li 2024** | China | 196 | 72(36.7) | 63.5(11.0) | Laparoscopic radical colorectal cancer resection | tDCS | Afternoon before and morning of surgery | CAM/CAM-ICU; 2 h after surgery to POD3 |
| **Zhou 2025** | China | 122 | 65(53.3) | 70.2(4.1) | Major abdominal surgery | rTMS | Immediately after extubation in PACU | CAM; POD1–3 |
| **Yang 2025** | China | 201 | 60(29.9) | 72.2(4.9)* | Major laparoscopic surgery >2 h | tDCS | 30 min before anaesthesia induction | CAM-ICU in PACU and CAM on ward; surgery day to POD3 |
| **Gao 2026** | China | 249 | 115(46.2) | 68.3(7.3)* | Elective non-cardiac surgery ≥2 h | rTMS | Day before and day of surgery | CAM/CAM-ICU; within 7 postoperative days |
| **Wei 2026** | China | 214 | 103(48.1) | 70.0(6.5) | Abdominal tumour surgery | tDCS | Afternoon before surgery and after extubation in PACU | RASS screen plus 3D-CAM/CAM-ICU; until discharge |
| **Zhang 2026** | China | 78 | 41(52.6) | 61.3(9.8) | Aortic or mitral valve surgery with cardiopulmonary bypass | cTBS | Reported course from day −1 through POD4 | CAM; first 5 postoperative days |
| **Antonenko 2026** | Germany | 180 | 76(42.2) | 73.0(6.0) | Mixed elective major surgery >60 min | α-tACS and tDCS | Postoperative PACU | 3D-CAM; up to 5 postoperative in-hospital days |
| **Chen 2026** | China | 89 | 38(42.7) | 76.5(3.0) | Hip fracture surgery | rTMS | Day −1, surgery day, and daily through POD7 | Paper-specific CAM thresholds; within 7 days |

**Abbreviations:** 3D-CAM, 3-Minute Diagnostic Interview for Confusion Assessment Method; α-tACS, alpha-frequency transcranial alternating current stimulation; CAM, Confusion Assessment Method; CAM-ICU, Confusion Assessment Method for the Intensive Care Unit; cTBS, continuous theta-burst stimulation; PACU, post-anaesthesia care unit; POD, postoperative delirium; RASS, Richmond Agitation-Sedation Scale; rTMS, repetitive transcranial magnetic stimulation; tDCS, transcranial direct current stimulation.
\* Age was calculated from group-level data reported in the original study.

### Postoperative delirium

Across nine trials, POD occurred in 72 of 750 participants allocated to active NIBS and 166 of 701 controls. Random-effects meta-analysis showed a lower risk of POD with NIBS (RR 0.41, 95% CI 0.31-0.56; tau^2^=0.0447; *I*^2^=24.7%; Q=10.62, 8 df, *P*=0.224; Figure 2). Eight trials had point estimates favouring active stimulation; the MODEST trial was the sole study with a point estimate favouring sham (RR 1.41, 95% CI 0.58-3.46).^18^

**Fig 2.**
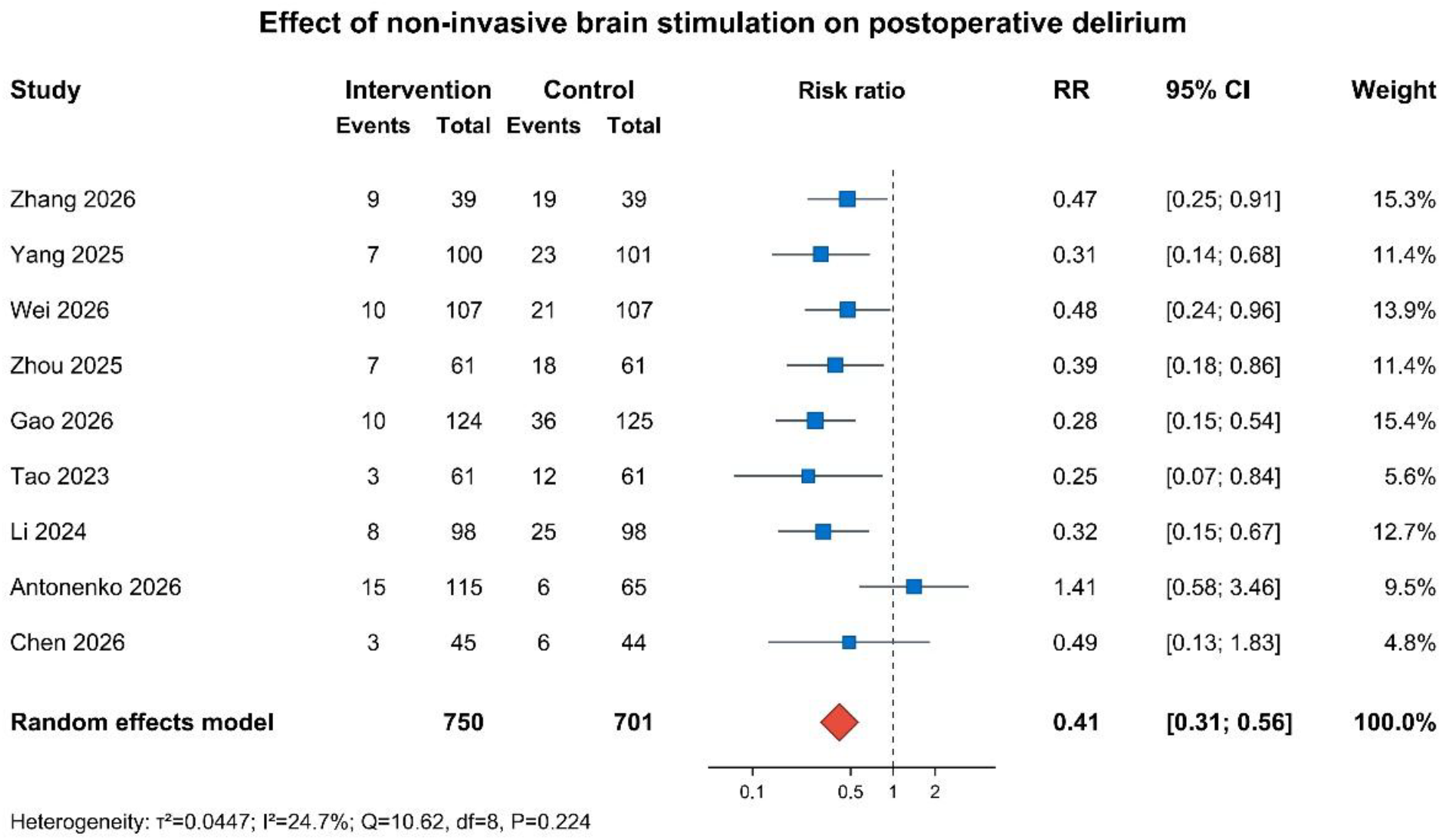
Forest plot of the effect of perioperative non-invasive brain stimulation on postoperative delirium. Risk ratios (RRs) and 95% confidence intervals (CIs) are presented for individual trials and the pooled estimate. The pooled effect was calculated using an inverse-variance random-effects model, with between-study variance estimated by restricted maximum likelihood. CI, confidence interval; RR, risk ratio.

### Delirium motor subtypes

Six studies (968 participants) reported delirium subtypes. NIBS was associated with lower risks of hypoactive delirium (RR 0.35, 95% CI 0.21-0.58; *I*^2^=0%) and hyperactive delirium (RR 0.31, 95% CI 0.18-0.52; *I*^2^=0%). The estimate for mixed delirium was compatible with both a clinically important reduction and no effect (RR 0.54, 95% CI 0.26-1.10; *I*^2^=0%; Figure 3).

**Fig 3.**
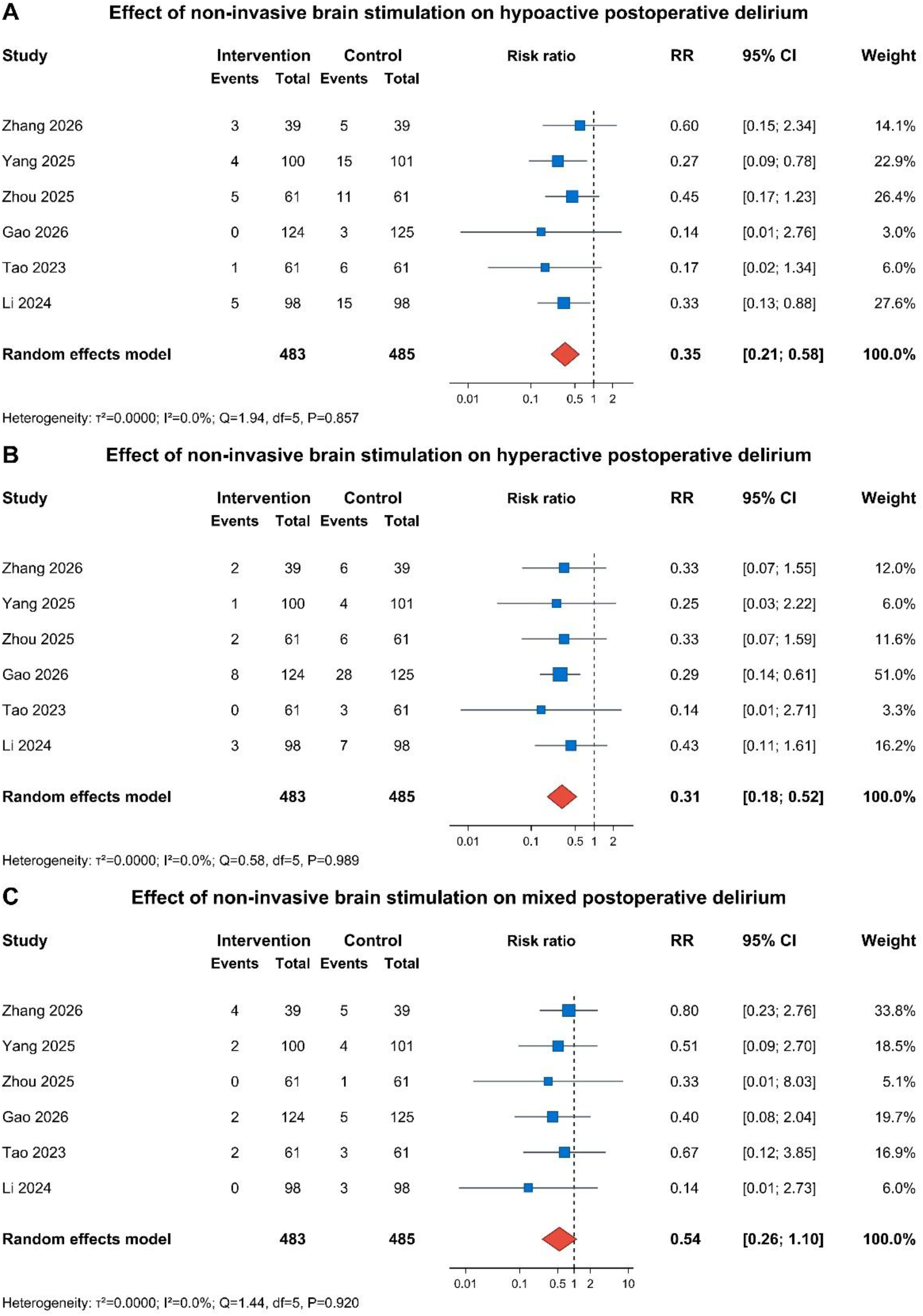
Forest plots of the effects of perioperative non-invasive brain stimulation on postoperative delirium motor subtypes, including hypoactive, hyperactive, and mixed delirium. Study-specific and pooled risk ratios (RRs) are presented with 95% confidence intervals (CIs) using random-effects meta-analysis. CI, confidence interval; RR, risk ratio.

#### Pain, anxiety, sleep, and hospital stay

Five trials (846 participants) contributed postoperative day 1 pain data. NIBS was associated with lower pain scores (MD −0.78, 95% CI −1.17 to −0.40), but heterogeneity was substantial (I^2^=91.4%). Four trials (597 participants) reported postoperative day 1 anxiety, with a pooled SMD of −0.48 (95% CI −0.85 to −0.11; *I*^2^=77.8%). Three trials (523 participants) reported postoperative day 1 sleep outcomes, with a pooled SMD of −0.86 (95% CI −1.33 to −0.39; *I*^2^=83.5%). Four trials (694 participants) reported length of hospital stay; there was no evidence of a reduction (MD 0.31 days, 95% CI −0.31 to 0.94; *I*^2^=0%; Figure 4).

**Fig 4.**
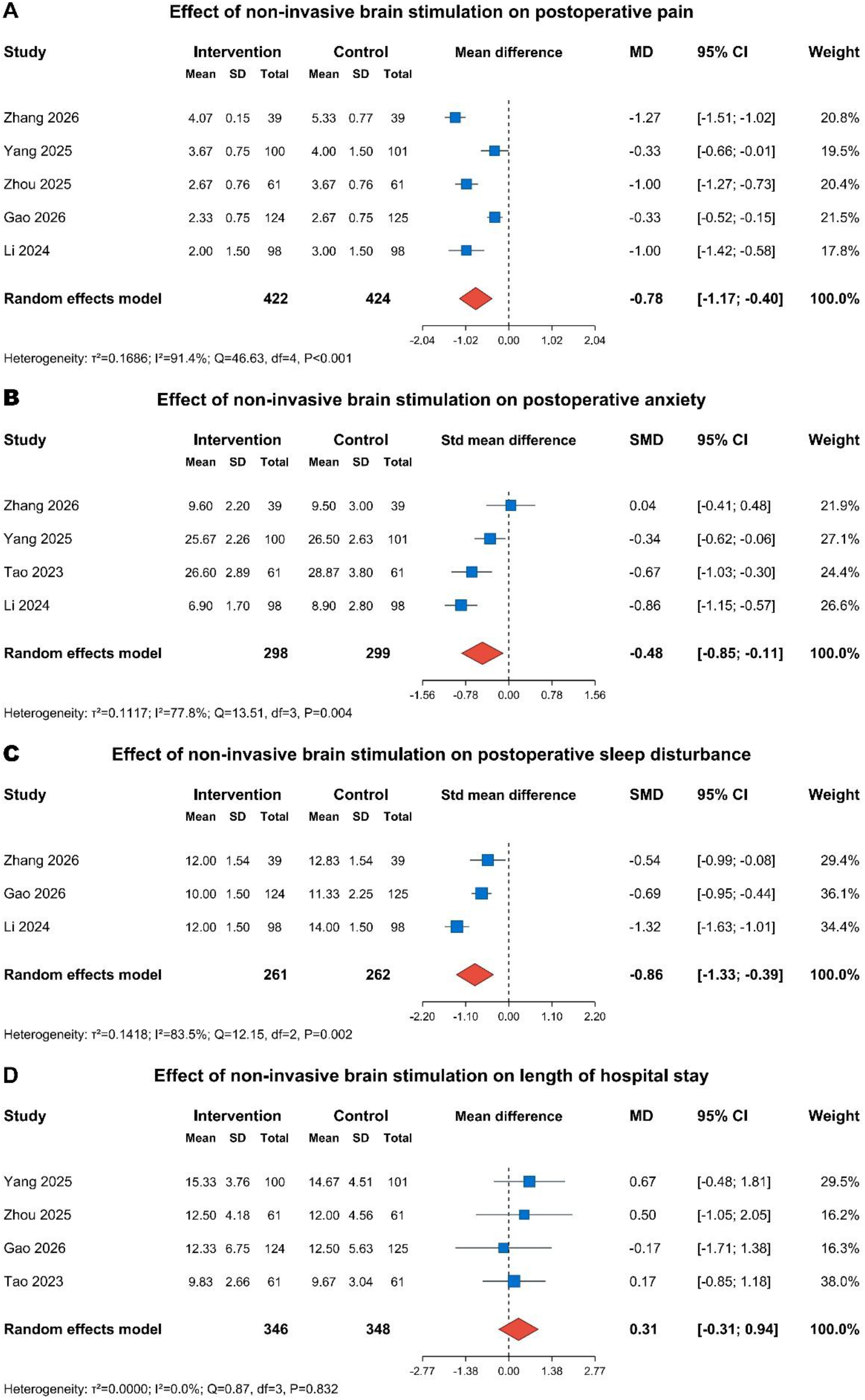
Forest plots of the effects of perioperative non-invasive brain stimulation on postoperative pain, anxiety, sleep disturbance, and length of hospital stay. Pain, anxiety, and sleep outcomes were assessed on postoperative day 1. Pooled effects are presented as mean differences (MDs) or standardised mean differences (SMDs), with 95% confidence intervals (CIs), using random-effects meta-analysis. CI, confidence interval; MD, mean difference; SMD, standardised mean difference.

### Risk of bias, sensitivity analyses, and stimulation modality

Two trials were judged at low overall risk of bias, four had some concerns, and three were at high overall risk (Supplementary Figure S1). High-risk judgments were driven principally by missing outcome data and outcome measurement, with additional concerns regarding selection of the reported result. Excluding the three high-risk studies retained a protective pooled estimate (RR 0.44, 95% CI 0.28-0.69; *I*^2^=49.7%; Supplementary Figure S5). Excluding Chen 2026 produced a similar result (RR 0.41, 95% CI 0.30-0.57; *I*^2^=33.7%; Supplementary Figure S3). Excluding MODEST strengthened the pooled estimate and removed observed heterogeneity (RR 0.37, 95% CI 0.28-0.48; *I*^2^=0%; Supplementary Figure S4), demonstrating that the recent discordant trial materially influenced between-study inconsistency. Modified Hartung-Knapp inference produced the same point estimate with a wider confidence interval (RR 0.41, 95% CI 0.29–0.60; Supplementary Figure S6).

In the modality subgroup analysis, magnetic stimulation (rTMS/cTBS; four studies) was associated with RR 0.38 (95% CI 0.26-0.56; *I*^2^=0%), whereas electrical stimulation (tDCS/tACS; five studies) was associated with RR 0.45 (95% CI 0.25-0.79; *I*^2^=54.8%). There was no statistical evidence of a subgroup interaction (Q=0.23, 1 df, *P*=0.635; Supplementary Figure S7). Given the small number of trials and within-modality heterogeneity, this analysis does not establish equivalence of magnetic and electrical approaches.

### Adverse events and certainty of evidence

Adverse events could not be meta-analysed. Most reports did not provide deduplicated participant-level counts for experiencing at least one adverse event, and several reported only selected categories or P values. One trial reported internally inconsistent total adverse-event counts, and the MODEST safety section did not state explicit event-report denominators for all comparisons (Supplementary Table S6). Consequently, absence of a pooled safety signal should not be interpreted as evidence of safety.

GRADE certainty was moderate for the primary POD outcome, downgraded one level for risk of bias (Table 2). Certainty was low for hypoactive, hyperactive, and mixed delirium because of risk of bias and imprecision. Evidence for pain, anxiety, and sleep outcomes was very low certainty because of risk of bias, substantial inconsistency, and imprecision. Length of stay was supported by low-certainty evidence. Safety evidence was of very low certainty because of risk of bias, very serious imprecision, and concerns about selective reporting; incomplete and non-standardised adverse-event reporting precluded quantitative synthesis. Visual inspection of the primary-outcome funnel plot was not considered sufficient to exclude small-study effects because only nine trials were available (Supplementary Figure S2).

**Table 2.**
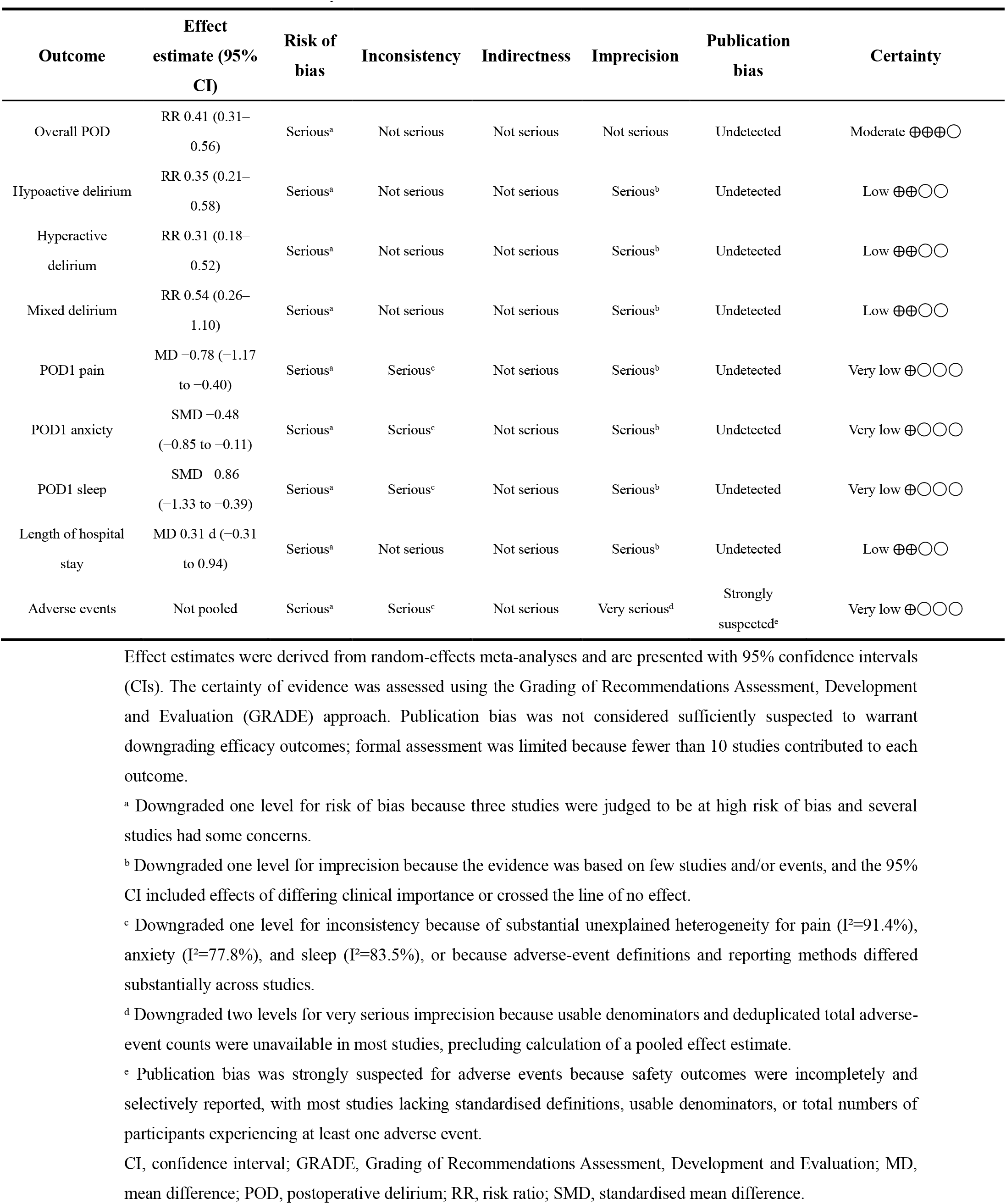
GRADE certainty of evidence for clinical outcomes.

| **Outcome** | **Effect estimate (95% CI)** | **Risk of bias** | **Inconsistency** | **Indirectness** | **Imprecision** | **Publication bias** | **Certainty** |
|---|---|---|---|---|---|---|---|
| Overall POD | RR 0.41 (0.31–0.56) | Serious<sup>a</sup> | Not serious | Not serious | Not serious | Undetected | Moderate ⊕⊕⊕◯ |
| Hypoactive delirium | RR 0.35 (0.21–0.58) | Serious<sup>a</sup> | Not serious | Not serious | Serious<sup>b</sup> | Undetected | Low ⊕⊕◯◯ |
| Hyperactive delirium | RR 0.31 (0.18–0.52) | Serious<sup>a</sup> | Not serious | Not serious | Serious<sup>b</sup> | Undetected | Low ⊕⊕◯◯ |
| Mixed delirium | RR 0.54 (0.26–1.10) | Serious<sup>a</sup> | Not serious | Not serious | Serious<sup>b</sup> | Undetected | Low ⊕⊕◯◯ |
| POD1 pain | MD −0.78 (−1.17 to −0.40) | Serious<sup>a</sup> | Serious<sup>c</sup> | Not serious | Serious<sup>b</sup> | Undetected | Very low ⊕◯◯◯ |
| POD1 anxiety | SMD −0.48 (−0.85 to −0.11) | Serious<sup>a</sup> | Serious<sup>c</sup> | Not serious | Serious<sup>b</sup> | Undetected | Very low ⊕◯◯◯ |
| POD1 sleep | SMD −0.86 (−1.33 to −0.39) | Serious<sup>a</sup> | Serious<sup>c</sup> | Not serious | Serious<sup>b</sup> | Undetected | Very low ⊕◯◯◯ |
| Length of hospital stay | MD 0.31 d (−0.31 to 0.94) | Serious<sup>a</sup> | Not serious | Not serious | Serious<sup>b</sup> | Undetected | Low ⊕⊕◯◯ |
| Adverse events | Not pooled | Serious<sup>a</sup> | Serious<sup>c</sup> | Not serious | Very serious<sup>d</sup> | Strongly suspected<sup>e</sup> | Very low ⊕◯◯◯ |

Effect estimates were derived from random-effects meta-analyses and are presented with 95% confidence intervals (CIs). The certainty of evidence was assessed using the Grading of Recommendations Assessment, Development and Evaluation (GRADE) approach. Publication bias was not considered sufficiently suspected to warrant downgrading efficacy outcomes; formal assessment was limited because fewer than 10 studies contributed to each outcome.
<sup>a</sup> Downgraded one level for risk of bias because three studies were judged to be at high risk of bias and several studies had some concerns.
<sup>b</sup> Downgraded one level for imprecision because the evidence was based on few studies and/or events, and the 95% CI included effects of differing clinical importance or crossed the line of no effect.
<sup>c</sup> Downgraded one level for inconsistency because of substantial unexplained heterogeneity for pain (I<sup>2</sup>=91.4%), anxiety (I<sup>2</sup>=77.8%), and sleep (I<sup>2</sup>=83.5%), or because adverse-event definitions and reporting methods differed substantially across studies.
<sup>d</sup> Downgraded two levels for very serious imprecision because usable denominators and deduplicated total adverse-event counts were unavailable in most studies, precluding calculation of a pooled effect estimate.
<sup>e</sup> Publication bias was strongly suspected for adverse events because safety outcomes were incompletely and selectively reported, with most studies lacking standardised definitions, usable denominators, or total numbers of participants experiencing at least one adverse event.
CI, confidence interval; GRADE, Grading of Recommendations Assessment, Development and Evaluation; MD, mean difference; POD, postoperative delirium; RR, risk ratio; SMD, standardised mean difference.

## Discussion

### Principal findings

In this systematic review of nine randomised trials, perioperative NIBS was associated with a substantially lower pooled risk of POD, with moderate certainty after downgrading for risk of bias. The direction of effect was robust to exclusion of high-risk studies, exclusion of the non-sham Chen comparison, and modified Hartung-Knapp inference. However, the pooled estimate masks an important evolution of the evidence base: eight predominantly Chinese trials favoured intervention, whereas the recent German MODEST trial did not show benefit and had a point estimate in the opposite direction. Removing MODEST reduced I^2^ to 0%, indicating that the principal source of statistical inconsistency was also the study that most directly challenges the reproducibility of the earlier signal.

Interpretation of the discordant evidence

The central interpretative problem is therefore not whether the conventional meta-analysis is statistically significant, but whether the pooled effect represents a transportable treatment effect. Similar tensions have arisen in other POD prevention literatures. In the BJA meta-analysis of processed EEG-guided anaesthesia, nine randomised trials showed a modest pooled benefit with important clinical heterogeneity; more recently, a BJA Bayesian synthesis of dexmedetomidine demonstrated that an apparently favourable pooled estimate became highly uncertain after accounting for publication bias and giving appropriate weight to the discordant DECADE trial.^8, 9^ Our findings have the same structural warning: early positive evidence and a later negative trial should not be reconciled simply by averaging them without examining why their results differ.

Several explanations are plausible. MODEST used a postoperative single-session electrical intervention, posterior parietal alpha-tACS and prefrontal tDCS, whereas positive trials varied from preoperative tDCS to postoperative rTMS and multi-session protocols.^10-18^ Baseline POD risk also differed. The sham event rate in MODEST was lower than in many earlier positive trials, limiting the absolute number of preventable events and potentially changing the balance between neural modulation and background delirium-prevention care. Differences in surgery, anaesthetic management, delirium ascertainment, stimulation target, and dose may also be effect modifiers. With only nine trials, these explanations cannot be separated reliably by meta-regression.

### Does a class effect of NIBS exist?

The absence of a statistically significant interaction between magnetic and electrical stimulation should not be interpreted as evidence that these modalities are equivalent. tDCS changes membrane polarisation, tACS seeks to entrain oscillatory activity, conventional rTMS uses repeated magnetic pulses to alter cortical excitability and network dynamics, and cTBS applies a patterned protocol with distinct physiological effects.^7^Even within a modality, target, intensity, frequency, number of sessions, timing relative to anaesthesia, and sham design differed markedly. The subgroup analysis therefore supports, at most, the observation that both broad modality classes contain positive trials; it does not validate a single shared biological effect size.

A mechanistic rationale nevertheless exists. Several included trials targeted the dorsolateral prefrontal cortex, a region involved in executive control and distributed attention networks, and the positive rTMS trial by Zhou and colleagues reported changes in brain-derived neurotrophic factor and neurofilament light compatible with a neuroplastic or neuroprotective pathway.^12^ Such biomarker findings are hypothesis-generating rather than confirmatory: mediation in a single trial cannot establish that the pooled clinical effect operates through the same mechanism across electrical and magnetic stimulation. Future trials should therefore prespecify mechanistic measures and relate target engagement to clinically adjudicated delirium rather than treating biomarker changes as validation of efficacy.

### Secondary outcomes and safety

Pain, anxiety, and sleep outcomes all favoured NIBS, but these estimates were accompanied by I^2^ values of 77.8-91.4% and very-low-certainty evidence. They should not be used to construct a secondary efficacy narrative around NIBS. Differences in measurement instruments, surgical populations, co-analgesia, timing, and stimulation protocols offer multiple non-exclusive explanations for the heterogeneity. Conversely, the absence of a difference in hospital stay suggests that the large relative reduction in POD did not translate into a measurable short-term resource-use benefit in the available trials, although the estimate was imprecise.

The safety evidence is weaker than the efficacy evidence. Standardised participant-level adverse-event reporting was absent in most trials, selected symptoms were often reported without a denominator for any adverse event, and category counts could not be summed without double counting. This prevents a quantitative estimate of overall tolerability and creates a risk of selective safety reporting. Established safety guidelines provide reassurance for commonly used tES and TMS parameters in controlled settings.^39, 40^ However, these external data cannot substitute for procedure-specific perioperative safety reporting in older and medically complex surgical patients.

### Strengths and limitations

This review has several strengths. The search covered major English-language and Chinese bibliographic databases and a clinical-trial registry, included only randomised trials, retained full study-level information on stimulation protocols and delirium ascertainment, used RoB 2 and GRADE, and tested the primary result with multiple sensitivity analyses. The analysis also avoids an important inferential error by treating the recent negative MODEST trial as evidence to be explained rather than as an outlier to be dismissed.

The limitations are substantial. First, eight of nine trials were conducted in China, restricting external validity and raising the possibility that site-specific perioperative pathways, patient selection, or publication processes contribute to the observed effect. Second, the evidence base remains small, with 1451 participants in the principal comparisons and relatively few POD events; subgroup analyses are consequently underpowered. Third, stimulation protocols were heterogeneous in target, intensity, frequency, timing, and session number. Fourth, POD ascertainment varied from CAM-based assessments over postoperative days 1-3 to 7-day or discharge-based windows, and one study used paper-specific CAM thresholds. Current perioperative guidance recommends repeated assessment with validated tools, so heterogeneity in ascertainment can alter measured incidence.^4^ Fifth, three trials were at high overall risk of bias and four had some concerns. Sixth, most trials were single-centre and relatively small, making exaggerated early effects and selective reporting plausible even though formal publication-bias tests were not informative with only nine studies.^37^Finally, adverse-event reporting was too incomplete to support a reliable safety estimate.

### Clinical and research implications

The pooled result is sufficiently large to justify further investigation but not sufficiently secure to justify routine perioperative NIBS for delirium prevention. A clinically persuasive next trial should be multicentre, sham-controlled, prospectively registered, and adequately powered for delirium as a primary outcome; use a harmonised validated assessment beginning in recovery and repeated for at least the first three postoperative days; prespecify adverse-event definitions and participant-level denominators; and standardise the stimulation target, dose, timing, and number of sessions. Independent replication outside China is particularly important. Trials should also prespecify whether the intervention is intended as a common NIBS class strategy or as a modality-specific treatment, because the present evidence does not establish interchangeability.

### Conclusions

Perioperative non-invasive brain stimulation was associated with a lower incidence of postoperative delirium in pooled randomised evidence, with moderate certainty for the primary outcome. The effect persisted in sensitivity analyses, but confidence in a broadly applicable treatment effect is limited by geographical concentration, small-study and risk-of-bias concerns, heterogeneous stimulation protocols, inconsistent safety reporting, and the discordant MODEST trial. Current evidence supports further definitive trials rather than routine clinical adoption.

### Declarations Ethics approval

Ethical approval was not required because this systematic review and meta-analysis used only published aggregate data and did not involve individual participant data.

## Data Availability

All data used in this systematic review and meta-analysis were derived from published studies. The extracted data supporting the findings of this study are provided in the manuscript and its supplementary materials.

## Registration

This systematic review was registered with PROSPERO (CRD420261522168).

## Author contributions

Conceptualisation: DZ, JZ, LD

Data curation: HW, HP, DL, AZ, YH, XL, JS, HZ

Methodology: HW, HP, DL, DZ, JZ, LD

Validation: HW, HP, DL, AZ, YH, XL, JS, HZ

Formal analysis: HW, HP, DL

Supervision: DZ, JZ, LD

Writing—original draft: HW, HP, DL

Writing—review and editing: all authors

Final approval of the manuscript and accountability for the work: all authors

## References

1 Inouye SK, Westendorp RG, Saczynski JS. Delirium in elderly people. Lancet 2014; 383: 911–22

2 Marcantonio ER. Delirium in Hospitalized Older Adults. N Engl J Med 2017; 377: 1456–663

3 Evered L, Silbert B, Knopman DS, et al. Recommendations for the nomenclature of cognitive change associated with anaesthesia and surgery-2018. Br J Anaesth 2018; 121: 1005–12

4 Aldecoa C, Bettelli G, Bilotta F, et al. Update of the European Society of Anaesthesiology and Intensive Care Medicine evidence-based and consensus-based guideline on postoperative delirium in adult patients. Eur J Anaesthesiol 2024; 41: 81–108

5 Hshieh TT, Yue J, Oh E, et al. Effectiveness of multicomponent nonpharmacological delirium interventions: a meta-analysis. JAMA Intern Med 2015; 175: 512–20

6 Luney M, Holdsworth L, Hagana A, et al. Effectiveness of drug interventions to prevent delirium after surgery for older adults: systematic review and network meta-analysis of randomised controlled trials. BMJ 2026; 392: e085539

7 Polanía R, Nitsche MA, Ruff CC. Studying and modifying brain function with non-invasive brain stimulation. Nat Neurosci 2018; 21: 174–87

8 Sumner M, Deng C, Evered L, et al. Processed electroencephalography-guided general anaesthesia to reduce postoperative delirium: a systematic review and meta-analysis. Br J Anaesth 2023; 130: e243–e53

9 Hunt T, Payne T, Brophy JM, et al. Perioperative dexmedetomidine for the prevention of postoperative delirium after cardiac surgery: a systematic review, Bayesian meta-analysis, and Bayesian re-analysis of the DECADE trial. Br J Anaesth 2025; 134: 1671–82

10 Tao M, Zhang S, Han Y, et al. Efficacy of transcranial direct current stimulation on postoperative delirium in elderly patients undergoing lower limb major arthroplasty: A randomized controlled trial. Brain Stimul 2023; 16: 88–96

11 Li C, Tao M, Chen D, et al. Transcranial Direct Current Stimulation for Anxiety During Laparoscopic Colorectal Cancer Surgery: A Randomized Clinical Trial. JAMA Netw Open 2024; 7: e246589

12 Zhou C, Gao YN, Qiao Q, et al. Efficacy of repetitive transcranial magnetic stimulation in preventing postoperative delirium in elderly patients undergoing major abdominal surgery: A randomized controlled trial. Brain Stimul 2025; 18: 52–60

13 Yang X, Wei YF, Ye CH, et al. Effect of preoperative transcranial direct current stimulation on postoperative delirium in elderly patients following laparoscopic surgery. World J Psychiatry 2025; 15: 107298

14 Gao WB, Wang WH, Zhou ST, et al. Efficacy of Repetitive Transcranial Magnetic Stimulation on Postoperative Delirium in Elderly Patients Undergoing Non-Cardiac Major Surgery: A Randomized Controlled Trial. Brain Behav 2026; 16: e71242

15 Wei S, Xiang X, Yao M, Ni H. Effect of transcranial direct current stimulation on perioperative neurocognitive disorder in elderly patients undergoing abdominal tumour surgery: A randomised controlled trial. Eur J Anaesthesiol 2026; 43: 627–30

16 Zhang J, Zhu Y, Yin Y, et al. Effect of Continuous Theta-Burst Stimulation on Postoperative Delirium in Older Patients Who Undergo Cardiac Surgery. Neuromodulation 2026

17 Chen B, Chen R, Guo Y, Wang L. Effects of ear holographic scraping therapy combined with repetitive transcranial magnetic stimulation on postoperative delirium and cognitive function in elderly patients undergoing hip fracture surgery]. Zhongguo Zhen Jiu 2026; 46: 1090–7

18 Antonenko D, Bublitz V, Leroy S, et al. Transcranial Electrical Stimulation and Delirium Among Postoperative Patients: A Randomized Clinical Trial. JAMA Netw Open 2026; 9: e2628284

19 Page MJ, McKenzie JE, Bossuyt PM, et al. The PRISMA 2020 statement: an updated guideline for reporting systematic reviews. BMJ 2021; 372: 71

20 Higgins JPT, Thomas J, Chandler J, et al., eds. Cochrane Handbook for Systematic Reviews of Interventions, version 6.5 (updated August 2024): Cochrane, 2024

21 Inouye SK, van Dyck CH, Alessi CA, Balkin S, Siegal AP, Horwitz RI. Clarifying confusion: the confusion assessment method. A new method for detection of delirium. Ann Intern Med 1990; 113: 941–8

22 Ely EW, Inouye SK, Bernard GR, et al. Delirium in mechanically ventilated patients: validity and reliability of the confusion assessment method for the intensive care unit (CAM-ICU). JAMA 2001; 286: 2703–10

23 Marcantonio ER, Ngo LH, O’Connor M, et al. 3D-CAM: derivation and validation of a 3-minute diagnostic interview for CAM-defined delirium: a cross-sectional diagnostic test study. Ann Intern Med 2014; 161: 554–61

24 Sterne JAC, Savović J, Page MJ, et al. RoB 2: a revised tool for assessing risk of bias in randomised trials. BMJ 2019; 366: 4898

25 McGuinness LA, Higgins JPT. Risk-of-bias VISualization (robvis): An R package and Shiny web app for visualizing risk-of-bias assessments. Res Synth Methods 2021; 12: 55–61

26 Balshem H, Helfand M, Schünemann HJ, et al. GRADE guidelines: 3. Rating the quality of evidence. J Clin Epidemiol 2011; 64: 401–6

27 Guyatt GH, Oxman AD, Vist G, et al. GRADE guidelines: 4. Rating the quality of evidence--study limitations (risk of bias). J Clin Epidemiol 2011; 64: 407–15

28 Guyatt GH, Oxman AD, Montori V, et al. GRADE guidelines: 5. Rating the quality of evidence--publication bias. J Clin Epidemiol 2011; 64: 1277–82

29 Guyatt GH, Oxman AD, Kunz R, et al. GRADE guidelines 6. Rating the quality of evidence--imprecision. J Clin Epidemiol 2011; 64: 1283–93

30 Guyatt GH, Oxman AD, Kunz R, et al. GRADE guidelines: 7. Rating the quality of evidence--inconsistency. J Clin Epidemiol 2011; 64: 1294–302

31 Guyatt GH, Oxman AD, Kunz R, et al. GRADE guidelines: 8. Rating the quality of evidence--indirectness. J Clin Epidemiol 2011; 64: 1303–10

32 Wan X, Wang W, Liu J, Tong T. Estimating the sample mean and standard deviation from the sample size, median, range and/or interquartile range. BMC Med Res Methodol 2014; 14: 135

33 Luo D, Wan X, Liu J, Tong T. Optimally estimating the sample mean from the sample size, median, mid-range, and/or mid-quartile range. Stat Methods Med Res 2018; 27: 1785–805

34 Higgins JP, Thompson SG, Deeks JJ, Altman DG. Measuring inconsistency in meta-analyses. BMJ 2003; 327: 557–60

35 Veroniki AA, Jackson D, Viechtbauer W, et al. Methods to estimate the between-study variance and its uncertainty in meta-analysis. Res Synth Methods 2016; 7: 55–79

36 IntHout J, Ioannidis JP, Borm GF. The Hartung-Knapp-Sidik-Jonkman method for random effects meta-analysis is straightforward and considerably outperforms the standard DerSimonian-Laird method. BMC Med Res Methodol 2014; 14: 25

37 Sterne JA, Sutton AJ, Ioannidis JP, et al. Recommendations for examining and interpreting funnel plot asymmetry in meta-analyses of randomised controlled trials. BMJ 2011; 343: d4002

38 Balduzzi S, Rücker G, Schwarzer G. How to perform a meta-analysis with R: a practical tutorial. Evid Based Ment Health 2019; 22: 153–60

39 Antal A, Bjekić J, Ganho-Ávila A, et al. Low intensity transcranial electric stimulation: Safety, ethical, legal regulatory and application guidelines (2017-2025: An update)-endorsed by the European Society for Brain Stimulation (ESBS) and by the International Federation for Clinical Neurophysiology (IFCN). Clin Neurophysiol 2026; 184: 2111436

40 Rossi S, Antal A, Bestmann S, et al. Safety and recommendations for TMS use in healthy subjects and patient populations, with updates on training, ethical and regulatory issues: Expert Guidelines. Clin Neurophysiol 2021; 132: 269–306

