## Supplementary Materials for "Perioperative non-invasive brain stimulation for the prevention of postoperative delirium: a systematic review and meta-analysis of randomised controlled trials"

**Supplementary Table S1. PRISMA 2020 checklist**

| Section and Topic | Item # | Checklist item | Location where item is reported |
|---|---|---|---|
| **TITLE** | | | |
| Title | 1 | Identify the report as a systematic review. | p 1 |
| **ABSTRACT** | | | |
| Abstract | 2 | See the PRISMA 2020 for Abstracts checklist. | p 2 |
| **INTRODUCTION** | | | |
| Rationale | 3 | Describe the rationale for the review in the context of existing knowledge. | pp 2–3 |
| Objectives | 4 | Provide an explicit statement of the objective(s) or question(s) the review addresses. | p 3 |
| **METHODS** | | | |
| Eligibility criteria | 5 | Specify the inclusion and exclusion criteria for the review and how studies were grouped for the syntheses. | pp 3–4 |
| Information sources | 6 | Specify all databases, registers, websites, organisations, reference lists and other sources searched or consulted to identify studies. Specify the date when each source was last searched or consulted. | p 4 |
| Search strategy | 7 | Present the full search strategies for all databases, registers and websites, including any filters and limits used. | Supplementary Table S2 |
| Selection process | 8 | Specify the methods used to decide whether a study met the inclusion criteria of the review, including how many reviewers screened each record and each report retrieved, whether they worked independently, and if applicable, details of automation tools used in the process. | p 4 |
| Data collection | 9 | Specify the methods used to collect data from reports, including how many reviewers collected data from each report, whether they worked independently, any processes for obtaining or confirming data from study investigators, and if | p 4 |

| Section and Topic | Item # | Checklist item | Location where item is reported |
|---|---|---|---|
| process | | applicable, details of automation tools used in the process. | |
| Data items | 10a | List and define all outcomes for which data were sought. Specify whether all results that were compatible with each outcome domain in each study were sought (e.g. for all measures, time points, analyses), and if not, the methods used to decide which results to collect. | p 4 |
| | 10b | List and define all other variables for which data were sought (e.g. participant and intervention characteristics, funding sources). Describe any assumptions made about any missing or unclear information. | p 4 |
| Study risk of bias assessment | 11 | Specify the methods used to assess risk of bias in the included studies, including details of the tool(s) used, how many reviewers assessed each study and whether they worked independently, and if applicable, details of automation tools used in the process. | pp 4–5 |
| Effect measures | 12 | Specify for each outcome the effect measure(s) (e.g. risk ratio, mean difference) used in the synthesis or presentation of results. | p 5 |
| Synthesis methods | 13a | Describe the processes used to decide which studies were eligible for each synthesis (e.g. tabulating the study intervention characteristics and comparing against the planned groups for each synthesis (item #5)). | pp 3–5 |
| | 13b | Describe any methods required to prepare the data for presentation or synthesis, such as handling of missing summary statistics, or data conversions. | p 5 |
| | 13c | Describe any methods used to tabulate or visually display results of individual studies and syntheses. | p 5; Figures 2–4 |
| | 13d | Describe any methods used to synthesize results and provide a rationale for the choice(s). If meta-analysis was performed, describe the model(s), method(s) to identify the presence and extent of statistical heterogeneity, and software package(s) used. | pp 5–6 |

| Section and Topic | Item # | Checklist item | Location where item is reported |
|---|---|---|---|
| | 13e | Describe any methods used to explore possible causes of heterogeneity among study results (e.g. subgroup analysis, meta-regression). | p 5 |
| | 13f | Describe any sensitivity analyses conducted to assess robustness of the synthesized results. | p 5 |
| Reporting bias assessment | 14 | Describe any methods used to assess risk of bias due to missing results in a synthesis (arising from reporting biases). | p 6 |
| Certainty assessment | 15 | Describe any methods used to assess certainty (or confidence) in the body of evidence for an outcome. | pp 4–5 |
| **RESULTS** | | | |
| Study selection | 16a | Describe the results of the search and selection process, from the number of records identified in the search to the number of studies included in the review, ideally using a flow diagram. | p 6; Figure 1 |
| | 16b | Cite studies that might appear to meet the inclusion criteria, but which were excluded, and explain why they were excluded. | Supplementary Table S3 |
| Study characteristics | 17 | Cite each included study and present its characteristics. | pp 6–7; Table 1 |
| Risk of bias in studies | 18 | Present assessments of risk of bias for each included study. | p 8; Supplementary Fig. S1 |
| Results of individual | 19 | For all outcomes, present, for each study: (a) summary statistics for each group (where appropriate) and (b) an effect estimate and its precision (e.g. confidence/credible interval), ideally using structured tables or plots. | pp 6–8; Figs 2–4; |

| Section and Topic | Item # | Checklist item | Location where item is reported |
|---|---|---|---|
| studies | | | Supplementary Figs S3–S7 |
| Results of syntheses | 20a | For each synthesis, briefly summarise the characteristics and risk of bias among contributing studies. | pp 6–8; Table 1; Supplementary Fig. S1 |
| | 20b | Present results of all statistical syntheses conducted. If meta-analysis was done, present for each the summary estimate and its precision (e.g. confidence/credible interval) and measures of statistical heterogeneity. If comparing groups, describe the direction of the effect. | pp 6–8; Figs 2–4; Supplementary Figs S3–S7 |
| | 20c | Present results of all investigations of possible causes of heterogeneity among study results. | p 8; Supplementary Fig. S7 |
| | 20d | Present results of all sensitivity analyses conducted to assess the robustness of the synthesized results. | p 8; Supplementary Figs S3–S6 |
| Reporting biases | 21 | Present assessments of risk of bias due to missing results (arising from reporting biases) for each synthesis assessed. | pp 6 and 8; Supplementary Fig. S2 |
| Certainty of evidence | 22 | Present assessments of certainty (or confidence) in the body of evidence for each outcome assessed. | pp 8–10; Table 2 |

| Section and Topic | Item # | Checklist item | Location where item is reported |
|---|---|---|---|
| **DISCUSSION** | | | |
| Discussion | 23a | Provide a general interpretation of the results in the context of other evidence. | pp 10–12 |
| | 23b | Discuss any limitations of the evidence included in the review. | p 11 |
| | 23c | Discuss any limitations of the review processes used. | p 11 |
| | 23d | Discuss implications of the results for practice, policy, and future research. | p 12 |
| **OTHER INFORMATION** | | | |
| Registration and protocol | 24a | Provide registration information for the review, including register name and registration number, or state that the review was not registered. | pp 2, 3, and 12 |
| | 24b | Indicate where the review protocol can be accessed, or state that a protocol was not prepared. | p 12 |
| | 24c | Describe and explain any amendments to information provided at registration or in the protocol. | p 12 |
| Support | 25 | Describe sources of financial or non-financial support for the review, and the role of the funders or sponsors in the review. | p 13 |
| Competing interests | 26 | Declare any competing interests of review authors. | p 13 |
| Availability of data, code and other materials | 27 | Report which of the following are publicly available and where they can be found: template data collection forms; data extracted from included studies; data used for all analyses; analytic code; any other materials used in the review. | p 13 |

**Supplementary Table S2. Search strategies for databases and trial registries**

All databases and trial registries were searched from inception to 30 September 2026

PubMed

| Search set | Search strategy | Records |
|---|---|---|
| #1 | ("Transcranial Magnetic Stimulation"[Mesh] OR "Transcranial Direct Current Stimulation"[Mesh] OR "transcranial magnetic stimulation"[Title/Abstract] OR "repetitive transcranial magnetic stimulation"[Title/Abstract] OR "repetitive transcranial magnet stimulation"[Title/Abstract] OR "theta burst stimulation"[Title/Abstract] OR "theta-burst stimulation"[Title/Abstract] OR "intermittent theta burst stimulation"[Title/Abstract] OR "continuous theta burst stimulation"[Title/Abstract] OR TMS[Title/Abstract] OR rTMS[Title/Abstract] OR TBS[Title/Abstract] OR iTBS[Title/Abstract] OR cTBS[Title/Abstract] OR "transcranial electrical stimulation"[Title/Abstract] OR "transcranial electric stimulation"[Title/Abstract] OR "transcranial electrical brain stimulation"[Title/Abstract] OR "transcranial electric brain stimulation"[Title/Abstract] OR "transcranial direct current stimulation"[Title/Abstract] OR "transcranial direct-current stimulation"[Title/Abstract] OR "transcranial alternating current stimulation"[Title/Abstract] OR "transcranial alternating-current stimulation"[Title/Abstract] OR "transcranial random noise stimulation"[Title/Abstract] OR "transcranial random-noise stimulation"[Title/Abstract] OR "transcranial pulsed current stimulation"[Title/Abstract] OR tES[Title/Abstract] OR tDCS[Title/Abstract] OR tACS[Title/Abstract] OR tRNS[Title/Abstract] OR tPCS[Title/Abstract] OR "non-invasive brain stimulation"[Title/Abstract] OR "noninvasive brain stimulation"[Title/Abstract] OR "non-invasive transcranial brain stimulation"[Title/Abstract] OR "noninvasive transcranial brain stimulation"[Title/Abstract] OR "non-invasive transcranial stimulation"[Title/Abstract] OR "noninvasive transcranial stimulation"[Title/Abstract] OR NIBS[Title/Abstract] OR NITBS[Title/Abstract] ) | 63,931 |
| #2 | ( "Delirium"[Mesh] OR delirium[Title/Abstract] OR delirious[Title/Abstract] OR "acute confusional state"[Title/Abstract] OR "acute confusional states"[Title/Abstract] OR "acute confusion"[Title/Abstract] OR "postoperative delirium"[Title/Abstract] OR "post-operative delirium"[Title/Abstract] OR "perioperative delirium"[Title/Abstract] OR "peri-operative delirium"[Title/Abstract] OR "perioperative neurocognitive disorder"[Title/Abstract] OR "perioperative neurocognitive disorders"[Title/Abstract] OR "peri-operative neurocognitive disorder"[Title/Abstract] OR "peri-operative neurocognitive disorders"[Title/Abstract] OR "perioperative neurocognitive dysfunction"[Title/Abstract] OR "peri-operative neurocognitive dysfunction"[Title/Abstract] OR "postoperative neurocognitive disorder"[Title/Abstract] OR "postoperative neurocognitive disorders"[Title/Abstract] OR "post-operative neurocognitive disorder"[Title/Abstract] OR "post-operative neurocognitive disorders"[Title/Abstract] OR "postoperative cognitive | 41,541 |

| Search set | Search strategy | Records |
|---|---|---|
| | dysfunction"[Title/Abstract] OR "post-operative cognitive dysfunction"[Title/Abstract] OR "postoperative cognitive decline"[Title/Abstract] OR "post-operative cognitive decline"[Title/Abstract] OR "postoperative cognitive impairment"[Title/Abstract] OR "post-operative cognitive impairment"[Title/Abstract] OR "delayed neurocognitive recovery"[Title/Abstract] OR PND[Title/Abstract] OR POCD[Title/Abstract]) | |
| #3 | #1 AND #2 | 62 |

Embase

| Search set | Search strategy | Records |
|---|---|---|
| #1 | ('transcranial magnetic stimulation'/exp OR 'transcranial direct current stimulation'/exp OR 'transcranial magnetic stimulation':ti,ab,kw OR 'repetitive transcranial magnetic stimulation':ti,ab,kw OR 'repetitive transcranial magnet stimulation':ti,ab,kw OR 'theta burst stimulation':ti,ab,kw OR 'theta-burst stimulation':ti,ab,kw OR 'intermittent theta burst stimulation':ti,ab,kw OR 'continuous theta burst stimulation':ti,ab,kw OR TMS:ti,ab,kw OR rTMS:ti,ab,kw OR TBS:ti,ab,kw OR iTBS:ti,ab,kw OR cTBS:ti,ab,kw OR 'transcranial electrical stimulation':ti,ab,kw OR 'transcranial electric stimulation':ti,ab,kw OR 'transcranial electrical brain stimulation':ti,ab,kw OR 'transcranial electric brain stimulation':ti,ab,kw OR 'transcranial direct current stimulation':ti,ab,kw OR 'transcranial direct-current stimulation':ti,ab,kw OR 'transcranial alternating current stimulation':ti,ab,kw OR 'transcranial alternating-current stimulation':ti,ab,kw OR 'transcranial random noise stimulation':ti,ab,kw OR 'transcranial random-noise stimulation':ti,ab,kw OR 'transcranial pulsed current stimulation':ti,ab,kw OR tES:ti,ab,kw OR tDCS:ti,ab,kw OR tACS:ti,ab,kw OR tRNS:ti,ab,kw OR tPCS:ti,ab,kw OR 'non-invasive brain stimulation':ti,ab,kw OR 'noninvasive brain stimulation':ti,ab,kw OR 'non-invasive transcranial brain stimulation':ti,ab,kw OR 'noninvasive transcranial brain stimulation':ti,ab,kw OR 'non-invasive transcranial stimulation':ti,ab,kw OR 'noninvasive transcranial stimulation':ti,ab,kw OR NIBS:ti,ab,kw OR NITBS:ti,ab,kw) | 6,498 |
| #2 | ('delirium'/exp OR delirium:ti,ab,kw OR delirious:ti,ab,kw OR 'acute confusional state':ti,ab,kw OR 'acute confusional states':ti,ab,kw OR 'acute confusion':ti,ab,kw OR 'postoperative delirium':ti,ab,kw OR 'post-operative delirium':ti,ab,kw OR 'perioperative delirium':ti,ab,kw OR 'peri-operative delirium':ti,ab,kw OR 'perioperative neurocognitive disorder':ti,ab,kw OR 'perioperative neurocognitive disorders':ti,ab,kw OR 'peri-operative neurocognitive disorder':ti,ab,kw OR 'peri-operative neurocognitive disorders':ti,ab,kw OR 'perioperative neurocognitive dysfunction':ti,ab,kw OR 'peri-operative neurocognitive dysfunction':ti,ab,kw OR 'postoperative neurocognitive disorder':ti,ab,kw OR 'postoperative neurocognitive disorders':ti,ab,kw OR 'post-operative neurocognitive disorder':ti,ab,kw OR 'post-operative neurocognitive disorders':ti,ab,kw OR 'postoperative cognitive | 3,413 |

| Search set | Search strategy | Records |
| --- | --- | --- |
| | dysfunction':ti,ab,kw OR 'post-operative cognitive dysfunction':ti,ab,kw OR 'postoperative cognitive decline':ti,ab,kw OR 'post-operative cognitive decline':ti,ab,kw OR 'postoperative cognitive impairment':ti,ab,kw OR 'post-operative cognitive impairment':ti,ab,kw OR 'delayed neurocognitive recovery':ti,ab,kw OR PND:ti,ab,kw OR POCD:ti,ab,kw ) | |
| #3 | #1 AND #2 | 28 |

CENTRAL

| Search set | Search strategy | Records |
| --- | --- | --- |
| #1 | MeSH descriptor: [Transcranial Magnetic Stimulation] explode all trees | 3,376 |
| #2 | MeSH descriptor: [Transcranial Direct Current Stimulation] explode all trees | 2,883 |
| #3 | ("transcranial magnetic stimulation" OR "repetitive transcranial magnetic stimulation" OR "repetitive transcranial magnet stimulation" OR "theta burst stimulation" OR "theta-burst stimulation" OR "intermittent theta burst stimulation" OR "continuous theta burst stimulation" OR TMS OR rTMS OR TBS OR iTBS OR cTBS OR "transcranial electrical stimulation" OR "transcranial electric stimulation" OR "transcranial electrical brain stimulation" OR "transcranial electric brain stimulation" OR "transcranial direct current stimulation" OR "transcranial direct-current stimulation" OR "transcranial alternating current stimulation" OR "transcranial alternating-current stimulation" OR "transcranial random noise stimulation" OR "transcranial random-noise stimulation" OR "transcranial pulsed current stimulation" OR tES OR tDCS OR tACS OR tRNS OR tPCS OR "non-invasive brain stimulation" OR "noninvasive brain stimulation" OR "non-invasive transcranial brain stimulation" OR "noninvasive transcranial brain stimulation" OR "non-invasive transcranial stimulation" OR "noninvasive transcranial stimulation" OR NIBS OR NITBS ):ti,ab,kw | 22,887 |
| #4 | #1 OR #2 OR #3 | 22,887 |
| #5 | MeSH descriptor: [Delirium] explode all trees | 2,052 |
| #6 | (delirium OR delirious OR "acute confusional state" OR "acute confusional states" OR "acute confusion" OR "postoperative delirium" OR "post-operative delirium" OR "perioperative delirium" OR "peri-operative delirium" OR "perioperative neurocognitive disorder" OR "perioperative neurocognitive disorders" OR "peri-operative neurocognitive disorder" OR "peri-operative neurocognitive disorders" OR "perioperative neurocognitive dysfunction" OR "peri-operative neurocognitive dysfunction" OR "postoperative neurocognitive disorder" OR "postoperative neurocognitive disorders" | 9,835 |

| Search set | Search strategy | Records |
|---|---|---|
| | OR "post-operative neurocognitive disorder" OR "post-operative neurocognitive disorders" OR "postoperative cognitive dysfunction" OR "post-operative cognitive dysfunction" OR "postoperative cognitive decline" OR "post-operative cognitive decline" OR "postoperative cognitive impairment" OR "post-operative cognitive impairment" OR "delayed neurocognitive recovery" OR PND OR POCD):ti,ab,kw | |
| #7 | #5 OR #6 | 9,835 |
| #8 | #4 AND #7 | 97 |

Web of Science

| Search set | Search strategy | Records |
|---|---|---|
| #1 | TS=( "transcranial magnetic stimulation" OR "repetitive transcranial magnetic stimulation" OR "repetitive transcranial magnet stimulation" OR "theta burst stimulation" OR "theta-burst stimulation" OR "intermittent theta burst stimulation" OR "continuous theta burst stimulation" OR TMS OR rTMS OR TBS OR iTBS OR cTBS OR "transcranial electrical stimulation" OR "transcranial electric stimulation" OR "transcranial electrical brain stimulation" OR "transcranial electric brain stimulation" OR "transcranial direct current stimulation" OR "transcranial direct-current stimulation" OR "transcranial alternating current stimulation" OR "transcranial alternating-current stimulation" OR "transcranial random noise stimulation" OR "transcranial random-noise stimulation" OR "transcranial pulsed current stimulation" OR tES OR tDCS OR tACS OR tRNS OR tPCS OR "non-invasive brain stimulation" OR "noninvasive brain stimulation" OR "non-invasive transcranial brain stimulation" OR "noninvasive transcranial brain stimulation" OR "non-invasive transcranial stimulation" OR "noninvasive transcranial stimulation" OR NIBS OR NITBS) | 106,214 |
| #2 | TS=(delirium OR delirious OR "acute confusional state" OR "acute confusional states" OR "acute confusion" OR "postoperative delirium" OR "post-operative delirium" OR "perioperative delirium" OR "peri-operative delirium" OR "perioperative neurocognitive disorder*" OR "peri-operative neurocognitive disorder*" OR "perioperative neurocognitive dysfunction" OR "peri-operative neurocognitive dysfunction" OR "postoperative neurocognitive disorder*" OR "post-operative neurocognitive disorder*" OR "postoperative cognitive dysfunction" OR "post-operative cognitive dysfunction" OR "postoperative cognitive decline" OR "post-operative cognitive decline" OR "postoperative cognitive impairment" OR "post-operative cognitive impairment" OR "delayed neurocognitive recovery" OR PND OR POCD) | 46,961 |
| #3 | #1 AND #2 | 67 |

CINAHL

| Search set | Search strategy | Records |
|---|---|---|
| #1 | ((MH "Transcranial Magnetic Stimulation+") OR (MH "Transcranial Direct Current Stimulation+") OR TI "transcranial magnetic stimulation" OR AB "transcranial magnetic stimulation" OR TI "repetitive transcranial magnetic stimulation" OR AB "repetitive transcranial magnetic stimulation" OR TI "repetitive transcranial magnet stimulation" OR AB "repetitive transcranial magnet stimulation" OR TI "theta burst stimulation" OR AB "theta burst stimulation" OR TI "theta-burst stimulation" OR AB "theta-burst stimulation" OR TI "intermittent theta burst stimulation" OR AB "intermittent theta burst stimulation" OR TI "continuous theta burst stimulation" OR AB "continuous theta burst stimulation" OR TI TMS OR AB TMS OR TI rTMS OR AB rTMS OR TI TBS OR AB TBS OR TI iTBS OR AB iTBS OR TI cTBS OR AB cTBS OR TI "transcranial electrical stimulation" OR AB "transcranial electrical stimulation" OR TI "transcranial electric stimulation" OR AB "transcranial electric stimulation" OR TI "transcranial electrical brain stimulation" OR AB "transcranial electrical brain stimulation" OR TI "transcranial electric brain stimulation" OR AB "transcranial electric brain stimulation" OR TI "transcranial direct current stimulation" OR AB "transcranial direct current stimulation" OR TI "transcranial direct-current stimulation" OR AB "transcranial direct-current stimulation" OR TI "transcranial alternating current stimulation" OR AB "transcranial alternating current stimulation" OR TI "transcranial alternating-current stimulation" OR AB "transcranial alternating-current stimulation" OR TI "transcranial random noise stimulation" OR AB "transcranial random noise stimulation" OR TI "transcranial random-noise stimulation" OR AB "transcranial random-noise stimulation" OR TI "transcranial pulsed current stimulation" OR AB "transcranial pulsed current stimulation" OR TI tES OR AB tES OR TI tDCS OR AB tDCS OR TI tACS OR AB tACS OR TI tRNS OR AB tRNS OR TI tPCS OR AB tPCS OR TI "non-invasive brain stimulation" OR AB "non-invasive brain stimulation" OR TI "noninvasive brain stimulation" OR AB "noninvasive brain stimulation" OR TI "non-invasive transcranial brain stimulation" OR AB "non-invasive transcranial brain stimulation" OR TI "noninvasive transcranial brain stimulation" OR AB "noninvasive transcranial brain stimulation" OR TI "non-invasive transcranial stimulation" OR AB "non-invasive transcranial stimulation" OR TI "noninvasive transcranial stimulation" OR AB "noninvasive transcranial stimulation" OR TI NIBS OR AB NIBS OR TI NITBS OR AB NITBS) | 13,407 |
| #2 | ((MH "Delirium+") OR TI delirium OR AB delirium OR TI delirious OR AB delirious OR TI "acute confusional state" OR AB "acute confusional state" OR TI "acute confusional states" OR AB "acute confusional states" OR TI "acute confusion" OR AB "acute confusion" OR TI "postoperative delirium" OR AB "postoperative delirium" OR TI "post-operative delirium" OR AB "post-operative delirium" OR TI "perioperative delirium" OR AB "perioperative delirium" OR TI "peri-operative delirium" OR AB "peri-operative delirium" OR TI "perioperative neurocognitive disorder" OR AB "perioperative neurocognitive disorder" OR TI "perioperative neurocognitive disorders" OR AB "perioperative neurocognitive disorders" OR TI "peri- | 16,903 |

| Search set | Search strategy | Records |
| --- | --- | --- |
| | operative neurocognitive disorder" OR AB "peri-operative neurocognitive disorder" OR TI "peri-operative neurocognitive disorders" OR AB "peri-operative neurocognitive disorders" OR TI "perioperative neurocognitive dysfunction" OR AB "perioperative neurocognitive dysfunction" OR TI "peri-operative neurocognitive dysfunction" OR AB "peri-operative neurocognitive dysfunction" OR TI "postoperative neurocognitive disorder" OR AB "postoperative neurocognitive disorder" OR TI "postoperative neurocognitive disorders" OR AB "postoperative neurocognitive disorders" OR TI "post-operative neurocognitive disorder" OR AB "post-operative neurocognitive disorder" OR TI "post-operative neurocognitive disorders" OR AB "post-operative neurocognitive disorders" OR TI "postoperative cognitive dysfunction" OR AB "postoperative cognitive dysfunction" OR TI "post-operative cognitive dysfunction" OR AB "post-operative cognitive dysfunction" OR TI "postoperative cognitive decline" OR AB "postoperative cognitive decline" OR TI "post-operative cognitive decline" OR AB "post-operative cognitive decline" OR TI "postoperative cognitive impairment" OR AB "postoperative cognitive impairment" OR TI "post-operative cognitive impairment" OR AB "post-operative cognitive impairment" OR TI "delayed neurocognitive recovery" OR AB "delayed neurocognitive recovery" OR TI PND OR AB PND OR TI POCD OR AB POCD) | |
| #3 | #1 AND #2 | 23 |

ClinicalTrials.gov

| Search set | Search strategy | Records |
| --- | --- | --- |
| #1 | "transcranial magnetic stimulation" OR "repetitive transcranial magnetic stimulation" OR "theta burst stimulation" OR TMS OR rTMS OR TBS OR iTBS OR cTBS OR "transcranial electrical stimulation" OR "transcranial direct current stimulation" OR "transcranial alternating current stimulation" OR "transcranial random noise stimulation" OR "transcranial pulsed current stimulation" OR tDCS OR tACS OR tRNS OR tPCS | 5,288 |
| #2 | delirium OR "postoperative delirium" OR "perioperative delirium" OR "perioperative neurocognitive disorder" OR "perioperative neurocognitive dysfunction" OR "postoperative neurocognitive disorder" OR "postoperative cognitive dysfunction" OR "delayed neurocognitive recovery" OR PND OR POCD | 1,937 |
| #3 | #1 AND #2 | 19 |

CNKI

| Search set | Search strategy | Records |
| --- | --- | --- |
| #1 | 经颅磁刺激+重复经颅磁刺激+重复性经颅磁刺激+经颅重复磁刺激+经颅重复性磁刺激+TMS+rTMS | 19,595 |
| #2 | θ 爆发刺激+θ 爆发式刺激+theta 爆发刺激+theta 节律爆发刺激+爆发式磁刺激+TBS+iTBS+cTBS | 2,413 |
| #3 | 经颅电刺激+经颅电流刺激+经颅直流电刺激+经颅直流刺激+tDCS+经颅交流电刺激+经颅交流刺激+tACS | 3,086 |
| #4 | 经颅随机噪声刺激+经颅随机噪音刺激+tRNS+经颅脉冲电流刺激+经颅脉冲电刺激+tPCS | 88 |
| #5 | 非侵入性脑刺激+非侵入性经颅脑刺激+非侵入性经颅刺激+无创脑刺激+无创经颅脑刺激+无创经颅刺激+NIBS+NITBS | 399 |
| #6 | #1 OR #2 OR #3 OR #4 OR #5 | 24,816 |
| #7 | 谵妄+术后谵妄+围术期谵妄+围手术期谵妄+急性意识混乱+急性意识障碍+急性混乱状态+delirium+POD | 58,051 |
| #8 | 围术期神经认知障碍+围手术期神经认知障碍+围术期神经认知功能障碍+围手术期神经认知功能障碍+术后神经认知障碍+术后神经认知功能障碍+PND | 1,705 |
| #9 | 术后认知功能障碍+术后认知障碍+术后认知功能下降+术后认知下降+延迟性神经认知恢复+延迟神经认知恢复+延迟性神经认知恢复障碍+POCD | 6,260 |
| #10 | #7 OR #8 OR #9 | 65,815 |
| #11 | #6 AND #10 | 11 |

Wanfang Data

| Search set | Search strategy | Records |
| --- | --- | --- |
| #1 | 主题:(经颅磁刺激 OR 重复经颅磁刺激 OR 重复性经颅磁刺激 OR 经颅重复磁刺激 OR TMS OR rTMS OR θ 爆发刺激 OR theta 爆发刺激 OR TBS OR iTBS OR cTBS OR 经颅电刺激 OR 经颅直流电刺激 OR tDCS OR 经颅交流电刺激 OR tACS OR 经颅随机噪声刺激 OR tRNS OR 经颅脉冲电刺激 OR tPCS OR 非侵入性脑刺激 OR 无创脑刺激 OR NIBS OR NITBS) AND 主题:(谵妄 OR 术后谵妄 OR 围术期谵妄 OR 围手术期谵妄 OR delirium OR "postoperative delirium" OR "perioperative delirium" OR POD OR 围术期神经认知障碍 OR 围手术期神经认知障碍 OR 术后神经认知障碍 OR 术后神经认知功能障碍 OR "perioperative neurocognitive disorder" OR "postoperative neurocognitive | 177 |

| Search set | Search strategy | Records |
|---|---|---|
| | disorder" OR PND OR 术后认知功能障碍 OR 术后认知障碍 OR "postoperative cognitive dysfunction" OR POCD OR 延迟性神经认知恢复 OR "delayed neurocognitive recovery") | |

SinoMed

| Search set | Search strategy | Records |
|---|---|---|
| #1 | (经颅磁刺激 OR 重复经颅磁刺激 OR 重复性经颅磁刺激 OR 经颅重复磁刺激 OR TMS OR rTMS OR θ 爆发刺激 OR theta 爆发刺激 OR TBS OR iTBS OR cTBS OR 经颅电刺激 OR 经颅直流电刺激 OR 经颅直流刺激 OR tDCS OR 经颅交流电刺激 OR 经颅交流刺激 OR tACS OR 经颅随机噪声刺激 OR tRNS OR 经颅脉冲电刺激 OR tPCS OR 非侵入性脑刺激 OR 无创脑刺激 OR NIBS OR NITBS) AND (谵妄 OR 术后谵妄 OR 围术期谵妄 OR 围手术期谵妄 OR delirium OR POD OR 围术期神经认知障碍 OR 围手术期神经认知障碍 OR 围术期神经认知功能障碍 OR 术后神经认知障碍 OR 术后神经认知功能障碍 OR 术后认知功能障碍 OR 术后认知障碍 OR PND OR POCD OR 延迟性神经认知恢复) | 24 |

**Abbreviations:** CENTRAL, Cochrane Central Register of Controlled Trials; CINAHL, Cumulative Index to Nursing and Allied Health Literature; CNKI, China National Knowledge Infrastructure.

**Supplementary Table S3. Reports and trial records excluded after full-text eligibility assessment, with reasons for exclusion**

| No. | registration | Title | Reason for exclusion |
|---|---|---|---|
| 1 | NCT04661904 | Effect of Continuous Theta Burst Stimulation (cTBS) on Postoperative Delirium | Trial registry only |
| 2 | NCT06678529 | Effect of Transcranial Direct Current Stimulation on Postoperative Delirium in Elderly Patients Undergoing Hip Fracture Surgery | Trial registry only |
| 3 | NCT07480044 | A Phase II Clinical Two-Stage Study of Transcranial Magnetic Stimulation in Preventing Postoperative Delirium in Elderly Patients After Urological Surgery | Trial registry only |
| 4 | CN-02748372 | Effect of transcranial alternating current stimulation on postoperative delirium in elderly patients undergoing non-cardiac surgery | Trial registry only |
| 5 | CN-02954320 | Effects of 40-Hz Transcranial Alternating Current Stimulation on Hypoactive Emergence Delirium and Postoperative Delirium in Elderly Noncardiac Surgical Patients: a Randomized Controlled Trial | Trial registry only |
| 6 | CN-02881399 | Effect of Perioperative re-Transcranial Electrical Stimulation on Postoperative Delirium in Patients Undergoing Off-Pump Coronary Artery Bypass Grafting: a Randomized Controlled Trial | Trial registry only |
| 7 | CN-02826763 | Transcranial Alternative Current Stimulation for postoperative delirium in elderly patients undergoing hip and knee replacement surgery: a randomized controlled trial | Trial registry only |
| 8 | CN-02868174 | Efficacy of intermittent theta burst stimulation (iTBS) on postoperative delirium in elderly patients undergoing non-cardiac major surgery: a prospective, double-blind, randomized controlled trial | Trial registry only |
| 9 | CN-02848730 | Efficacy of Repetitive Transcranial Magnetic Stimulation on Postoperative Delirium in Elderly Patients undergoing Major Lower Limb Arthroplasty: a Randomized Controlled Clinical Trial | Trial registry only |
| 10 | CN-02836001 | The effect of transcranial magnetic stimulation on postoperative delirium in elderly patients undergoing lower limb arthroplasty: a prospective, double-blind, randomized controlled trial | Trial registry only |
| 11 | CN-03065337 | Synergistic Effects of Dexmedetomidine and Transcranial Direct Current Stimulation in Preventing Postoperative Delirium Among Elderly Patients: a Prospective RCT | Trial registry only |
| 12 | CN-02984609 | The Effect of Transcranial Electrical Stimulation on Postoperative Delirium in Cardiac Surgery Patients: a Randomized Controlled Clinical Trial | Trial registry only |

| | | | |
|---|---|---|---|
| 13 | CN-02976230 | Effects of Perioperative Transcranial Alternating Current Stimulation on Postoperative Delirium in Elderly Patients | Trial registry only |
| 14 | CN-03025962 | The effect of transcranial alternating current stimulation on postoperative delirium incidence in elderly patients undergoing non cardiac surgery | Trial registry only |
| 15 | CN-03015230 | Efficacy of Repetitive Transcranial Magnetic Stimulation for the Prevention of Postoperative Delirium in Elderly Patients Undergoing Total Hip Arthroplasty | Trial registry only |
| 16 | CN-03049413 | Effects of Dorsolateral Prefrontal rTMS on Postoperative Delirium in Elderly Patients Undergoing Cardiac Surgery and Exploration of Its Neural Mechanisms: a Multimodal Neuroimaging Clinical Study Based on EEG and MRI | Trial registry only |
| 17 | CN-02732847 | Investigating the effect of TDCS on delirium | Trial registry only |
| 18 | CN-02652470 | The Effect of Transcranial Electrical Stimulation on Emergence Agaitation Following General Anesthesia: a Prospectitvie Study | Insufficient outcome reporting |
| 19 | CN-02785687 | Effect of transcranial direct current stimulation on emergence agitation in patients undergoing major abdominal surgery: a prospective randomized controlled clinical trial | Insufficient outcome reporting |
| 20 | CN-02902952 | Effects of transcranial direct current stimulation in perioperative depression in patients with breast cancer surgery | Insufficient outcome reporting |
| 21 | CN-02817774 | The effect of repetitive transcranial magnetic stimulation on perioperative anxiety in female patients: a single center, double-blind, randomized controlled study | Insufficient outcome reporting |
| 22 | CN-03015492 | The efficacy of transcranial alternating current stimulation on the quality of postoperative recovery in patients undergoing lumbar fusion surgery | Insufficient outcome reporting |
| 23 | CN-03060266 | Effect of Repetitive Transcranial Magnetic Stimulation on Perioperative Anxiety in Patients Undergoing Thoracoscopic Lung Resection: a Prospective, Single-Center, Randomized Controlled Study | Insufficient outcome reporting |
| 24 | CN-02981665 | The Effect of Repetitive Transcranial Magnetic Stimulation on Postoperative Recovery Quality in Patients Undergoing Major Abdominal Surgery: a Single-Center, Prospective, Randomized Controlled Study | Insufficient outcome reporting |

| | | | |
|---|---|---|---|
| 25 | NCT05668559 | The Effects of rTMS on Postoperative Cognitive Function in Patients After Cardiac Surgery | Ineligible outcome |
| 26 | CN-02853255 | Impact of repetitive transcranial magnetic stimulation on postoperative neurocognitive recovery in older patients with preoperative cognitive impairment | Ineligible outcome |
| 27 | CN-02905702 | A Study on Repetitive Transcranial Magnetic Stimulation (rTMS) for Anxiety and Sleep Disorders in Perioperative Patients | Ineligible outcome |
| 28 | CN-02934271 | Effect of repetitive transcranial magnetic stimulation on postoperative sleep disorders in patients undergoing open spinal surgery: a single-center, prospective, randomized controlled study | Ineligible outcome |
| 29 | NCT07100197 | EEG-TMS Intervening Against Postoperative Delirium in Elderly Patients: A Randomized Clinical Trial | POD treatment rather than prevention |
| 30 | NCT07537725 | Transcranial Magnetic Stimulation for Postoperative Delirium After Cardiac Surgery: A Prospective, Single-Center, Randomized Double-Blind Controlled Study | POD treatment rather than prevention |
| 31 | NCT06392919 | Effects of Perioperative Repetitive Transcranial Magnetic Stimulation on Postoperative Delirium, Postoperative Cognitive Dysfunction, and Chronic Post-surgical Pain in Elderly Surgical Patients | Duplicate report |
| 32 | CN-03018108 | Dexmedetomidine-esketamine and tDCS for Prevention of Neurocognitive Complications After Surgery | Ineligible study design |
| 33 | 10.3389/fpsyt.2026.1907287 | Prevention of postoperative delirium with transcranial alternating current stimulation: study protocol for a randomised controlled trial | Study protocol only |

**Supplementary Table S4. Detailed stimulation parameters of the included studies**

| Study | Target / montage | Intensity / frequency | Session protocol | Timing / number of sessions | Sham / comparator |
|---|---|---|---|---|---|
| Tao 2023 | Anode: left DLPFC; cathode: right orbitofrontal area | 2 mA direct current | 20 min; 30-s ramp-up and 30-s ramp-down | Single session immediately after extubation in the PACU | Sham tDCS: ramp-up for 30 s and ramp-down for 30 s, without sustained 2-mA stimulation |
| Li 2024 | Anode: left DLPFC; cathode: right DLPFC | 2 mA direct current | 20 min; 30-s ramp-up and 30-s ramp-down | Two sessions: afternoon before surgery and morning of surgery | Sham tDCS: 30-s ramp-up and 30-s ramp-down, without sustained 2-mA stimulation |
| Zhou 2025 | Left DLPFC; EEG 10-20 based positioning | 10 Hz; 100% RMT | 50 pulses/train; 25-s inter-train interval; 40 trains; 2000 pulses; 20 min | Single session after extubation in the PACU | Sham rTMS: same RMT assessment and coil position, but without active stimulation |
| Yang 2025 | Anode: F3 (left DLPFC); cathode: 1-2 cm above the right eyebrow | 2 mA direct current | 20 min; 30-s ramp-up and 30-s ramp-down | Single session initiated 30 min before anaesthesia induction | Sham tDCS: no sustained current during the 19-min middle phase; identical ramp-up/ramp-down |
| Wei 2026 | Anode: left DLPFC; cathode: right orbitofrontal area | 2 mA direct current | 20 min; 30-s ramp-up and 30-s ramp-down | Two sessions: afternoon before surgery and after extubation in the PACU | Sham tDCS: identical setup with current only during the 30-s ramp-up and ramp-down phases |
| Gao 2026 | Left DLPFC; EEG 10-20 based positioning | 10 Hz; 110% RMT | 2-s trains; 20-s inter-train interval; 1080 pulses; 20 min | Two sessions: day before surgery and day of surgery, separated by ≥ 12h | Sham rTMS: coil tilted approximately 90 degrees to the scalp to mimic acoustic sensation |

| Study | Target / montage | Intensity / frequency | Session protocol | Timing / number of sessions | Sham / comparator |
|---|---|---|---|---|---|
| Zhang 2026 | Posterior cingulate cortex; neuron-navigation and EEG 10-20 based positioning | Individual RMT-based intensity; exact percentage NR; bursts at 5 Hz with 3 pulses at 50 Hz per burst | 600 pulses; approximately 40 s per session | Five sessions: once daily from 1 day before surgery through POD3 | Sham cTBS: coil tilted 45 degrees and positioned about 5 cm lateral to F3/F4 over the temporalis muscle |
| Antonenko 2026 (MODEST) | P3 and P4; posterior parietal regions | 9.5 Hz; 2 mA (zero-to-peak) | 20 min; 10-s fade-in and 10-s fade-out | Single postoperative session in the PACU | Shared sham: current for 30 s with either the tACS or tDCS setup, plus 10-s ramp-up and ramp-down |
| Antonenko 2026 (MODEST) | Anode: F3 (left DLPFC); cathode: Fp2 | 2 mA direct current | 20 min; 10-s fade-in and 10-s fade-out | Single postoperative session in the PACU | Shared sham: current for 30 s with either the tACS or tDCS setup, plus 10-s ramp-up and ramp-down |
| Chen 2026 | Left DLPFC | 10 Hz; 90% motor threshold | 2-s trains; 20-s inter-train interval; 500 pulses/session | Once daily on the day before surgery, day of surgery, and postoperative days 1-7; 9 sessions total | Comparator: conventional multimodal delirium prevention; no sham stimulation |

Abbreviations: cTBS, continuous theta-burst stimulation; DLPFC, dorsolateral prefrontal cortex; EEG, electroencephalography; NR, not reported; PACU, post-anaesthesia care unit; PCC, posterior cingulate cortex; RMT, resting motor threshold; rTMS, repetitive transcranial magnetic stimulation; tACS, transcranial alternating current stimulation; tDCS, transcranial direct current stimulation.

Note: For the MODEST trial, tACS and tDCS were separate active arms that shared a single sham group. For Chen 2026, the rTMS-alone arm versus conventional care is shown because this comparison was used in the meta-analysis.

**Supplementary Table S5. Outcome measures and assessment instruments used in the included studies**

| Study | Overall POD | POD subtype | Postoperative pain | Anxiety | Sleep | Length of hospital stay |
|---|---|---|---|---|---|---|
| Tao 2023 | CAM / CAM-ICU | RASS | NRS | SAS | — | Days |
| Li 2024 | CAM / CAM-ICU | Clinical classification* | NRS | HADS-A | PSQI | — |
| Zhou 2025 | CAM | RASS | NRS | —† | — | Days |
| Yang 2025 | CAM-ICU / CAM | Clinical classification* | NRS | SAS | — | Days |
| Gao 2026 | CAM / CAM-ICU | RASS | NRS | —† | SDRS | Days |
| Wei 2026 | RASS screening + 3D-CAM / CAM-ICU | — | — | — | — | — |
| Zhang 2026 | CAM | RASS | NRS | Hamilton Anxiety Rating Scale–Anxiety subscale | PSQI | — |
| Antonenko 2026 | 3D-CAM | RASS | NRS | — | — | — |
| Chen 2026 | CAM-based score thresholds‡ | — | VAS§ | — | — | Days |

**Abbreviations:** CAM, Confusion Assessment Method; CAM-ICU, Confusion Assessment Method for the Intensive Care Unit; 3D-CAM, 3-Minute Diagnostic Interview for CAM-defined Delirium; HADS-A, Hospital Anxiety and Depression Scale–Anxiety subscale; NRS, Numeric Rating Scale; PSQI, Pittsburgh Sleep Quality Index; RASS, Richmond Agitation-Sedation Scale; SAS, Self-Rating Anxiety Scale; SDRS, Sleep Dysfunction Rating Scale; VAS, Visual Analogue Scale; POD, postoperative delirium.

*POD subtype was reported as hypoactive, hyperactive, or mixed, but no specific classification scale was explicitly reported.

†Anxiety was reported in the original study, but the available measurement timepoint did not meet the prespecified POD1 meta-analysis timepoint.

‡Chen 2026 used paper-specific CAM score thresholds rather than the standard CAM diagnostic algorithm.

§VAS data were reported in Chen 2026 but were not included in the pooled POD1 pain analysis.

**Supplementary Table S6. Study-level reporting of adverse events**

| Study | Analysis population, intervention/control | Any adverse event | Serious adverse event | Reported events and limitations | Meta-analysis decision |
|---|---|---|---|---|---|
| **Tao 2023** | 61/61 | Deduplicated totals not reported | Not reported separately | Several postoperative events and an overall comparison P value were reported, but it was unclear whether events occurred in different participants. | Not pooled |
| **Li 2024** | 98/98 | Totals not reported | Not reported | Skin tingling was reported in 11/98 versus 3/98; this did not represent the incidence of any adverse event. | Not pooled |
| **Zhou 2025** | 61/61 | Deduplicated totals not reported | Not reported | Individual event categories and an overall comparison P value were reported; category counts could not be summed without double-counting participants. | Not pooled |
| **Yang 2025** | 100/101 | Totals not reported | Not reported | Stimulation-site pain and other postoperative complications were described, but a complete participant-level total was unavailable. | Not pooled |
| **Gao 2026** | 124/125 | 24/124 vs 14/125 | No serious events reported | The text and component counts supported 24 versus 14, but the total column in the article was internally inconsistent. | Not pooled because of internal inconsistency |
| **Wei 2026** | 107/107 | Totals not reported | 0/107 vs 0/107 | No serious adverse events occurred; usable participant counts for non-serious skin irritation were not provided. | Not pooled |
| **Zhang 2026** | 39/39 | Deduplicated totals not reported | Not reported | Headache/dizziness, nausea/vomiting, stimulation-site pain, fever, and other categories were reported, but the same participant could contribute to more than one category. | Not pooled |

| Study | Analysis population, intervention/control | Any adverse event | Serious adverse event | Reported events and limitations | Meta-analysis decision |
|---|---|---|---|---|---|
| **Antonenko 2026 (MODEST)** | 115 active/65 sham | Not estimable; 27 active vs 8 sham events were reported, but participant-level event denominators were not explicit | 0 vs 0 | Any non-serious self-reported event was recorded as 10 tACS plus 17 tDCS events versus 8 sham events; the safety-section denominators were not explicitly stated. | Not pooled |
| **Chen 2026** | 45/44 | Totals not reported | 0/45 vs 0/44 for serious infection or bleeding | No uniform total for general adverse events was reported for the rTMS-alone versus conventional-prevention comparison. | Not pooled |

The analysis population column gives the denominators used for the principal intervention-control comparison where available. Counts for different adverse-event categories were not summed because the same participant could have experienced more than one event. A pooled effect estimate for any adverse event was therefore not calculated. tACS, transcranial alternating current stimulation; tDCS, transcranial direct current stimulation; rTMS, repetitive transcranial magnetic stimulation.

**Supplementary Figure S1. Risk-of-bias assessment of the included randomised controlled trials using the Cochrane Risk of Bias 2 (RoB 2) tool.**

Risk of bias assessment for overall postoperative delirium

| Study | D1 Randomization | D2 Deviations | D3 Missing data | D4 Measurement | D5 Reporting | Overall |
|---|---|---|---|---|---|---|
| Tao 2023 | + | + | + | + | + | + |
| Li 2024 | + | + | + | + | + | + |
| Zhou 2025 | + | + | - | + | ! | - |
| Yang 2025 | + | + | - | + | ! | - |
| Gao 2026 | + | + | ! | + | ! | ! |
| Wei 2026 | ! | + | + | + | ! | ! |
| Zhang 2026 | + | + | ! | + | ! | ! |
| Antonenko 2026 | + | + | ! | + | + | ! |
| Chen 2026 | + | ! | + | - | ! | - |

+ Low risk    ! Some concerns    - High risk

**Supplementary Figure S2. Funnel plot for the effect of perioperative non-invasive brain stimulation on postoperative delirium**

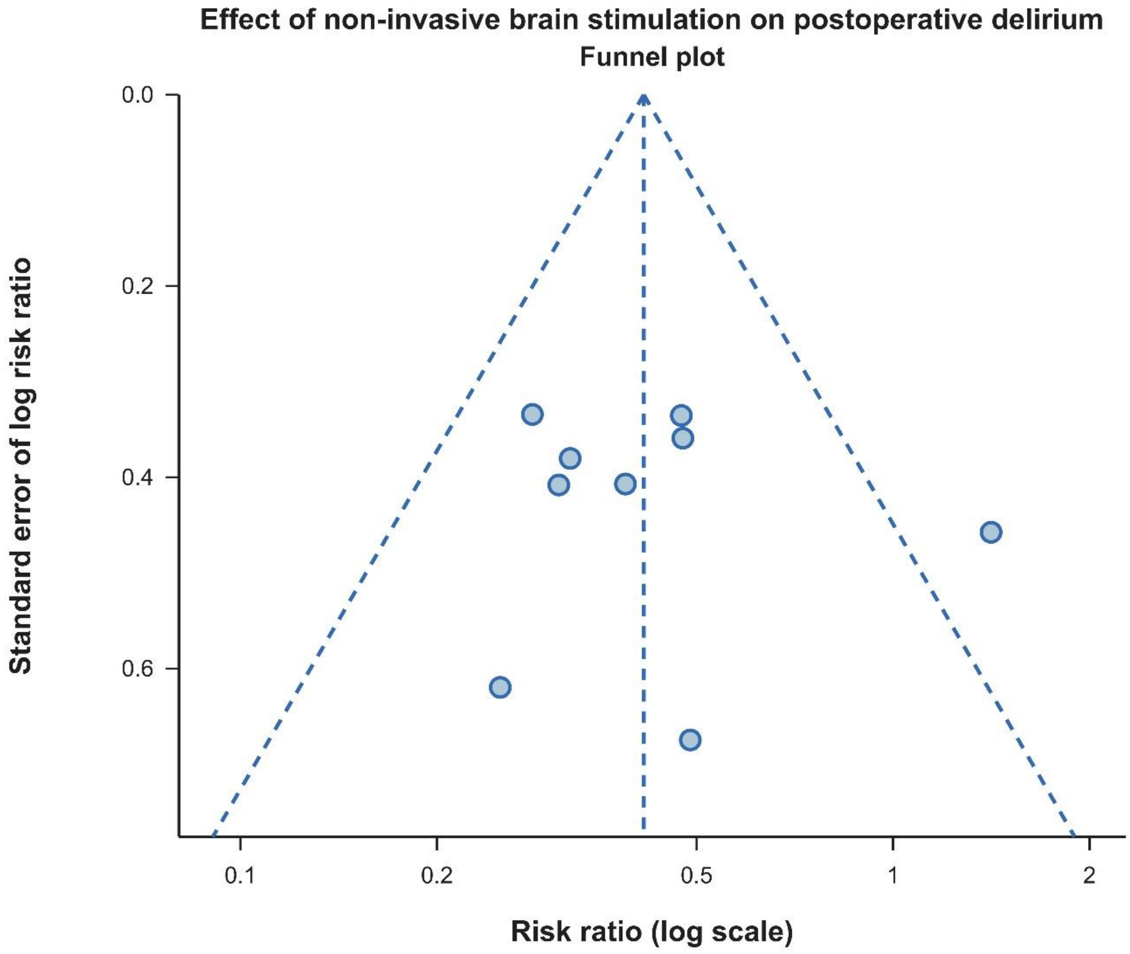

**Supplementary Figure S3. Sensitivity analysis for postoperative delirium after excluding Chen 2026**

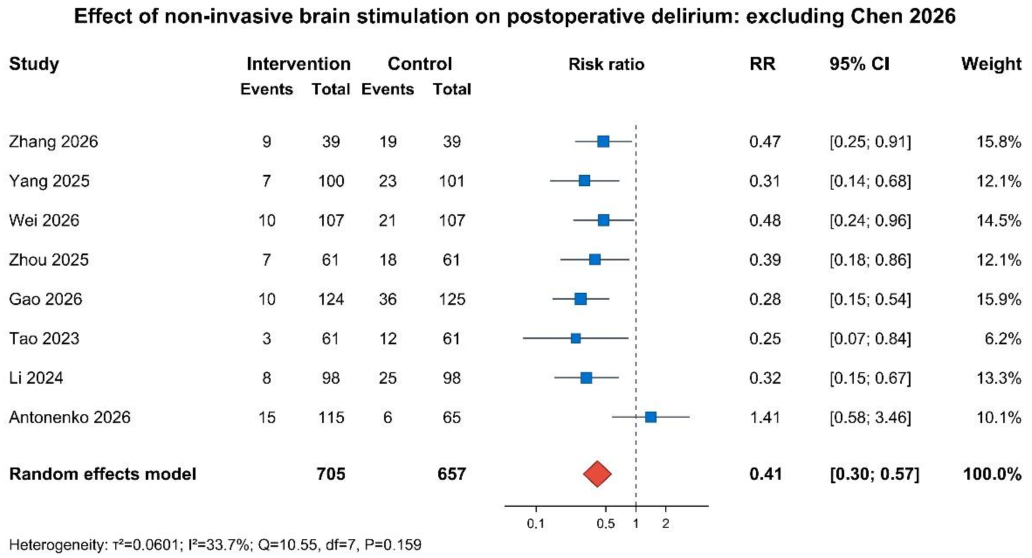

**Supplementary Figure S4. Sensitivity analysis for postoperative delirium after excluding the MODEST trial**

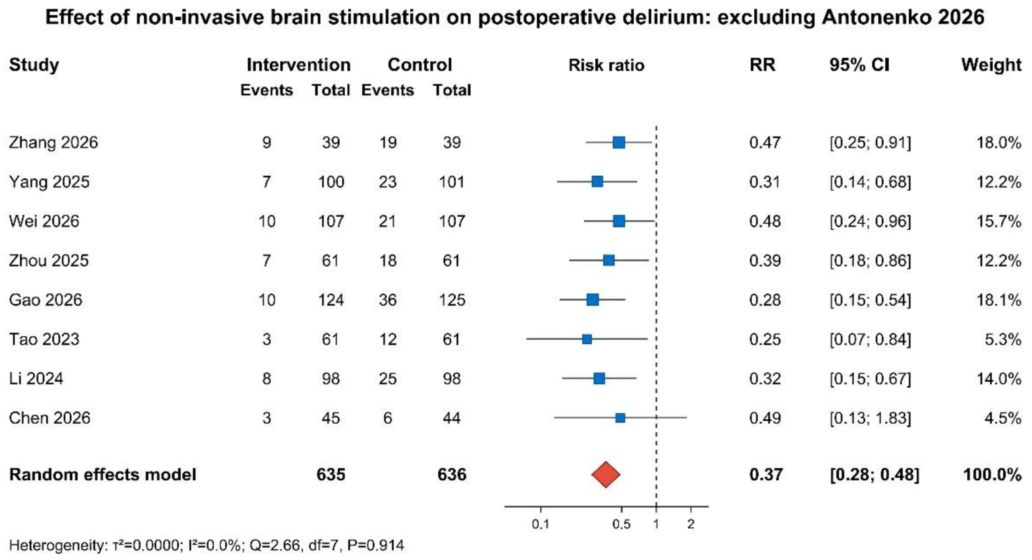

**Supplementary Figure S5. Sensitivity analysis for postoperative delirium after excluding studies at high risk of bias**

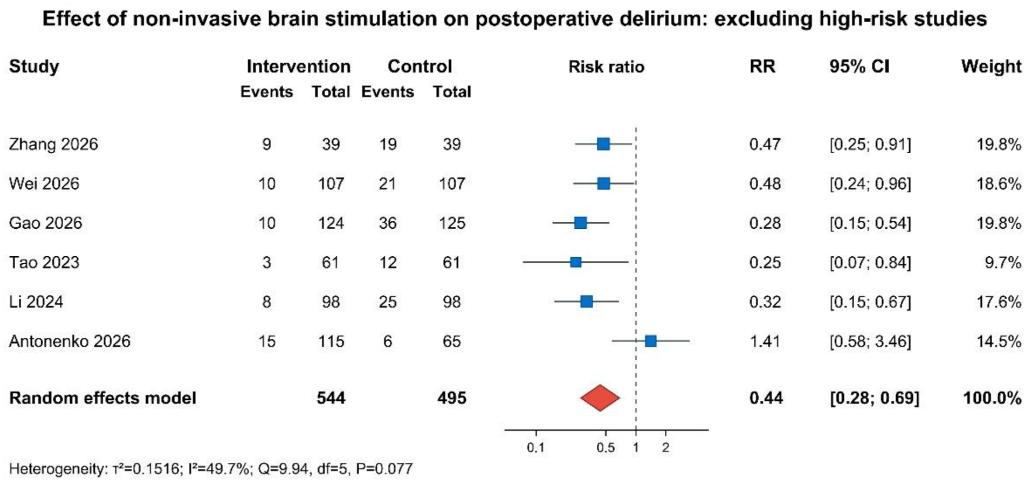

**Supplementary Figure S6. Modified Hartung–Knapp sensitivity analysis for postoperative delirium**

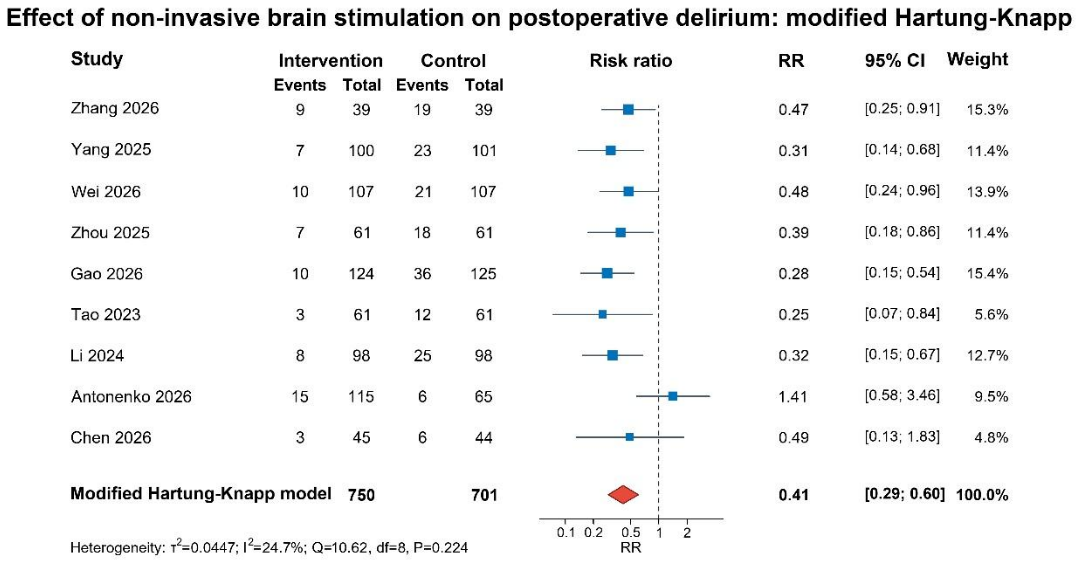

**Supplementary Figure S7. Subgroup analysis of postoperative delirium by stimulation modality**

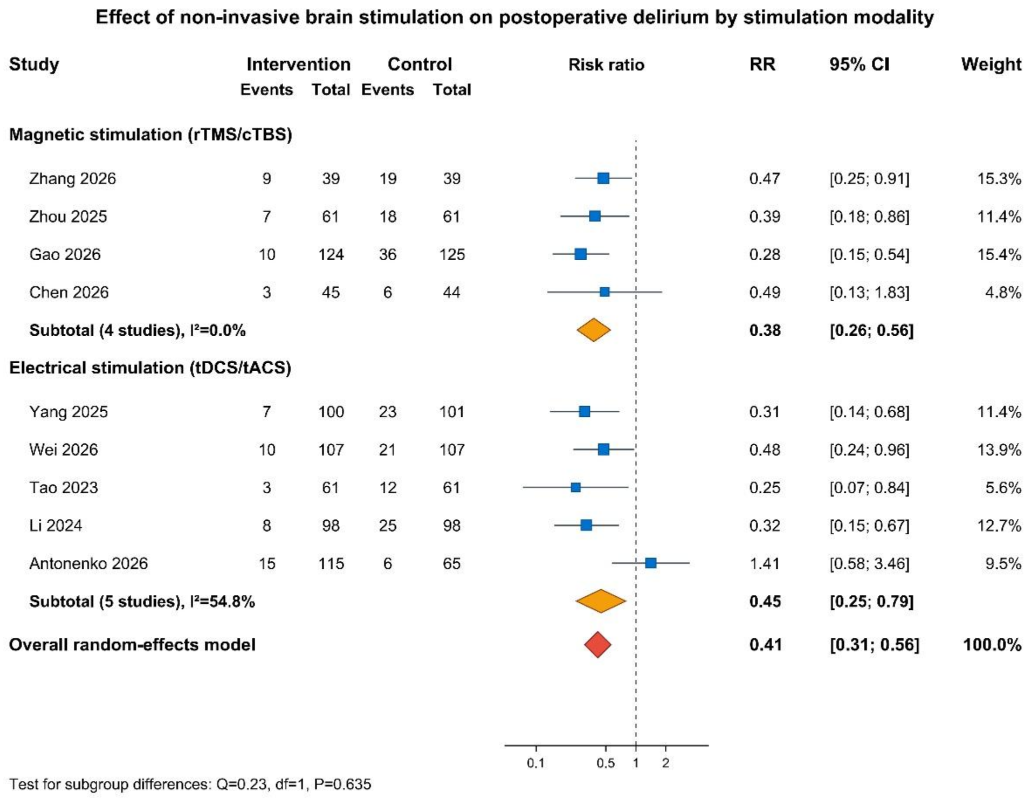
